# *APOE*-*ε*4 and *BIN1* increase risk of Alzheimer’s disease pathology but not specifically of Lewy body pathology

**DOI:** 10.1101/2023.04.21.23288938

**Authors:** Seth Talyansky, Yann Le Guen, Nandita Kasireddy, Michael E. Belloy, Michael D. Greicius

## Abstract

Lewy body (LB) pathology commonly occurs in individuals with Alzheimer’s disease (AD) pathology. However, it remains unclear which genetic risk factors underlie AD pathology, LB pathology, or AD-LB co-pathology. Notably, whether *APOE*-*ε*4 affects risk of LB pathology independently from AD pathology is controversial. We adapted criteria from the literature to classify 4,985 subjects from the National Alzheimer’s Coordinating Center (NACC) and the Rush University Medical Center as AD-LB co-pathology (AD^+^LB^+^), sole AD pathology (AD^+^LB^−^), sole LB pathology (AD^−^LB^+^), or no pathology (AD^−^LB^−^). We performed a meta-analysis of a genome-wide association study (GWAS) per subpopulation (NACC/Rush) for each disease phenotype compared to the control group (AD^−^LB^−^), and compared the AD^+^LB^+^ to AD^+^LB^−^ groups. *APOE*-*ε*4 was significantly associated with risk of AD^+^LB^−^ and AD^+^LB^+^ compared to AD^−^LB^−^. However, *APOE*-*ε*4 was not associated with risk of AD^−^LB^+^ compared to AD^−^LB^−^ or risk of AD^+^LB^+^ compared to AD^+^LB^−^. Associations at the *BIN1* locus exhibited qualitatively similar results. These results suggest that *APOE*-*ε*4 is a risk factor for AD pathology, but not for LB pathology when decoupled from AD pathology. The same holds for *BIN1* risk variants. These findings, in the largest AD-LB neuropathology GWAS to date, distinguish the genetic risk factors for sole and dual AD-LB pathology phenotypes. Our GWAS meta-analysis summary statistics, derived from phenotypes based on postmortem pathologic evaluation, may provide more accurate disease-specific polygenic risk scores compared to GWAS based on clinical diagnoses, which are likely confounded by undetected dual pathology and clinical misdiagnoses of dementia type.

## Introduction

Alzheimer’s disease (AD) pathology has been the focus of many studies, but Lewy body (LB) pathology has received less attention. In individuals with AD, LB pathology frequently co-occurs with AD pathology, while LB pathology alone or accompanied by limited AD pathology is characteristic of Parkinson’s disease (PD) and LB dementia [30, 53, 49, 54]. Genetic variants associated with AD pathology, LB pathology, and/or AD-LB co-pathology remain to be distinguished. Characterizing each set of risk factors and any potential overlap will help clarify the etiology of AD and LB pathology. AD pathology is found in 19–67% of older individuals at autopsy, depending on the population, the individual’s age, and the diagnostic criteria used [44]. LB pathology is observed in 6–39% of older individuals, but in 60% of individuals with AD pathology [44, 30, 34]. Positive classification for LB pathology requires, at a minimum, the presence of α-synuclein-bearing Lewy bodies in the brainstem, with further extension of LB pathology into the limbic system associated with the clinical diagnosis of dementia [38]. Positive classification for AD pathology requires the presence of tau neurofibrillary tangles (NFTs) in the limbic system along with amyloid-β core neuritic plaques in the cortex [8, 39]. Lewy bodies and NFTs spread to the cortex late in the progression of either pathology [38, 8]. Braak staging is the primary scheme used to classify NFT progression, while the Consortium to Establish a Registry for Alzheimer’s Disease (CERAD) scoring is the primary scheme used to classify neuritic plaque density. AD has traditionally been defined pathologically as Braak stage IV or higher, and at least moderate CERAD [17], although some studies have used less stringent criteria (Braak stage III or higher, and at least sparse CERAD) [28]. Thal phasing of amyloid-β non-neuritic plaques [51], based on another staging devised by Braak [9], has more recently been used as a third essential metric to classify AD [11]. Of the five Thal phases, only the last two, in which plaques are present in the brainstem and cerebellum, are specific to dementia patients [17].

Clinical diagnoses of AD and LB dementia are challenging and error-prone in comparison to the gold standard of a pathologic diagnosis [19]. However, most study participants have only been clinically diagnosed due to the scarcity of postmortem pathologically confirmed data. In a study of 919 autopsied individuals comparing clinical diagnosis of AD to pathological diagnosis, the diagnosis of clinically probable AD had an 83% positive predictive value (PPV) for pathological criteria of Braak state III or higher and moderate/high CERAD [1]. This study was conducted before AD biomarkers like spinal fluid amyloid and tau levels, or amyloid and tau PET scans, were more commonly used, so the PPV of the clinical diagnosis is now likely higher than 83%; however, it remains imperfect. The PPV for a clinical diagnosis of probable LB dementia against the pathologic diagnosis is also around 80% [21, 50, 45]. In general, it has been difficult to clinically distinguish between AD without Lewy bodies, AD with Lewy bodies, and LB dementia [25]. Additionally, individuals who may have advanced pathology, but mild symptoms are frequently misdiagnosed clinically or missing from clinical datasets altogether because they do not seek medical attention.

Motor function and neuropsychiatric and cognitive symptoms have been suggested as diagnostic clues of AD pathology, LB pathology, or co-pathology [14, 48]. Moreover, developing AD pathology biomarkers such as assays of amyloid-β, tau, and phosphorylated tau levels in the cerebrospinal fluid or blood plasma has been valuable in closing the gap between diagnosis during life and pathologic AD diagnosis [6, 32]. LB pathology biomarkers, including promising assays of α-synuclein aggregates in the cerebrospinal fluid, are similarly improving the diagnosis of LB dementia [47, 43, 36]. Still, because LB biomarkers have been developed more recently, most existing genetic datasets consist of only clinically diagnosed subjects. Ultimately, as a histological and molecular endophenotype, pathologic diagnosis offers the most reliable insights into the genetic drivers of disease.

Previous research has produced contrasting and somewhat ambiguous findings on the genetic risk loci for AD and LB pathology. This could be because most studies include only clinically assessed subjects or have relatively few pathologically assessed subjects. Importantly, most prior studies on AD and LB pathology, even with pathologic confirmation, do not stratify subjects into distinct groups for sole AD pathology (AD^+^LB^−^), LB pathology (AD^−^LB^+^), co-pathology (AD^+^LB^+^), and neither pathology (AD^−^LB^−^), making the results difficult to accurately interpret. It is well known that the *ε*4 allele of the *Apolipoprotein E* (*APOE*) gene is the strongest common genetic risk factor for AD [4, 31]. However, various studies have reported that *APOE*-*ε*4 is also associated with increased risk of sole LB pathology (AD^−^LB^+^) [52, 18], LB dementia [5, 2, 24, 48, 13], or increased risk of AD-LB co-pathology (AD^+^LB^+^) in AD individuals [14]. Walker and Richardson (2023) found that *APOE*-*ε*4 was associated with AD, LB, or limbic-predominant age-related TDP-43 encephalopathy pathology as well as with the presence of multiple of these pathologies [54]. This suggests that *APOE*-*ε*4 could be associated with AD^−^LB^+^ pathology.

How *APOE*-*ε*4 affects the severity of LB pathology has also been investigated. Studies reported that α-synuclein pathology mouse models expressing *APOE*-*ε*4 develop more extensive inclusions [22, 16]. In humans, LB pathology was found to be more severe among *APOE*-*ε*4 carriers independent of AD pathology severity [23], as well as among *APOE*-*ε*4-carrying AD^−^LB^+^ subjects relative to non-carriers [18, 22, 57]. However, when Kaivola *et al.* (2022) categorized pathologically confirmed cases from the cohort in [13] by not only LB but also AD pathology status, *APOE*-*ε*4 was not associated with risk of AD^−^LB^+^ pathology [28]. Robinson *et al.* (2018) found that *APOE*-*ε*4 was associated with cortical LB co-pathology (cortical LB pathology accompanied by an amyloidopathy, tauopathy, or TDP-43 proteinopathy) compared to sole LB pathology; however, *APOE*-*ε*4 was not associated with AD-LB co-pathology compared to sole AD pathology [46]. Furthermore, Dickson *et al.* (2018) found that *APOE*-*ε*4 was not associated with more severe LB pathology in individuals with moderate or high AD pathology [18]. It, therefore, remains unclear whether *APOE*-*ε*4 in fact increases risk of AD^−^LB^+^ pathology.

Importantly, beyond *APOE*-*ε*4, there may be other pathology-specific genetic risk loci yet to be identified. Along this line, it is relevant to note that removing individuals that are not pathologically evaluated from study cohorts has been shown to reduce noise in genome-wide association studies (GWAS) and to improve polygenic risk score analyses of AD [15, 20, 19]. These observations emphasize the need for novel GWAS of AD/LB pathology to better characterize the genetic architecture of these complex dementias.

To this end, we assembled a preliminary cohort of 5,254 individuals with genetic data and autopsy-confirmed AD and LB pathology status, the largest such cohort to date. We adapted criteria from the literature to categorize these individuals as AD^+^LB^+^, AD^+^LB^−^, AD^−^LB^+^, or AD^−^LB^−^, yielding 1,072 AD^+^LB^+^, 2,492 AD^+^LB^−^, 158 AD^−^LB^+^, and 1,263 AD^−^LB^−^ individuals in our study cohort (total N = 4,985). We compared each disease category to controls by performing separate GWAS meta-analyses. We also compared AD^+^LB^+^ pathology to AD^+^LB^−^ pathology in another analysis.

## Materials and methods

### Study cohort

We analyzed data from individuals from the National Alzheimer’s Coordinating Center (NACC) and Rush University Medical Center databases who were evaluated postmortem for both AD and LB pathology. We excluded NACC individuals who were classified as having Lewy bodies in the olfactory bulb or in an “unspecified” region (individuals for whom the NACCLEWY parameter was equal to 4). We also excluded individuals missing sex or age-at-death information. In total, our preliminary cohort comprised 5,254 individuals before classification according to AD and LB pathology status. This cohort was distinct from that analyzed in [13], the largest genetic study of LB dementia (which included subjects without pathology verification), and [28], the largest previous genetic study of subjects categorized by both AD and LB pathology status.

### Pathological criteria

We classified individuals as having both AD and LB pathology (AD^+^LB^+^), AD pathology only (AD^+^LB^−^), LB pathology only (AD^−^LB^+^), or neither pathology (AD^−^LB^−^) (**Fig. 1**). Individuals who could not be classified using our criteria were excluded (**Fig. 1c**). In sensitivity analyses, we applied the pathology criteria from [52] and [28] to our preliminary cohort **(Fig 1. a–b).** Criteria were set as follows for LB pathology.

- LB^+^ pathology were individuals with Lewy bodies spread to the limbic system or cortex, as in [28] and [52].
- LB^−^ pathology were individuals with no Lewy bodies or Lewy bodies limited to the brainstem, as in [28], but not in [52], which excluded individuals with brainstem-limited Lewy bodies.
- Some gray zones, representing rare subcategories with unclassified individuals, are defined based on Braak stage and CERAD score below.

**Figure 1.**
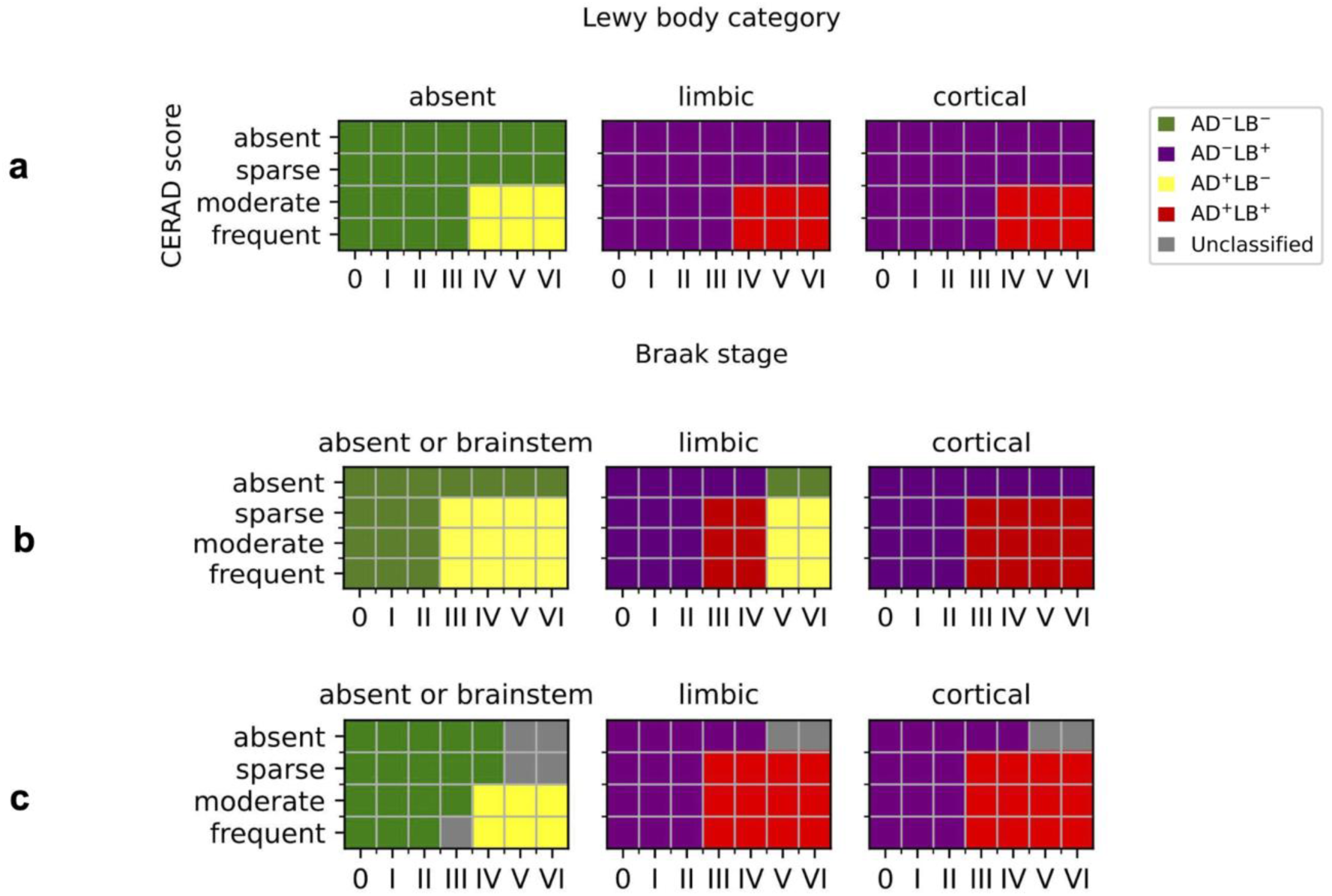
Schemes used to classify individuals. **a.** Criteria from Tsuang *et al.* (2013) [52]. **b.** Criteria from Kaivola *et al.* (2022) [28]. **c.** Criteria in the present study.

Criteria for AD pathology were less straightforward because of borderline subcategories and possible confounding with other pathologies. In agreement with [28] and [52] (**Fig. 1**),

- AD^+^ pathology included individuals with Braak stage IV or higher and CERAD score moderate or frequent.
- AD^−^ pathology included individuals with Braak stage II or lower, or Braak stage III/IV and CERAD score absent.

Other subcategories were largely classified differently between [28] and [52], and we settled on the following criteria. First, we defined three gray zones of unclassified individuals corresponding to rare pathologic profiles (N = 269 total).

- Individuals who had Braak stage V/VI and CERAD score absent, regardless of LB category, were not classified for the reason of likely having a rare tauopathy distinct from AD [41].
- Similarly, individuals with absent or brainstem Lewy bodies (LB^−^) who had Braak stage V/VI and CERAD score sparse were not classified.
- Braak stage III and CERAD score frequent in LB^−^ individuals were insufficient for classification as AD^+^, but too high for a confident classification as AD^−^. Ultimately, our goal was to obtain a clean control (AD^−^LB^−^) group.

Second, certain borderline subcategories were classified differently (AD^+^ or AD^−^) depending on LB category.

- Individuals with absent or brainstem Lewy bodies (LB^−^) were AD^−^ for Braak stage III and CERAD score sparse or moderate; or Braak stage IV and CERAD score sparse.
- Individuals with limbic or cortical Lewy bodies (LB^+^) were AD^+^ for Braak stage III or higher and CERAD score sparse; or Braak stage III and CERAD score moderate or frequent.

Using our criteria (**Fig. 1c**), we categorized our preliminary cohort into 1,072 AD^+^LB^+^, 2,495 AD^+^LB^−^, 158 AD^−^LB^+^, and 1,263 AD^−^LB^−^ individuals; these 4,985 individuals formed our study cohort (**Table 1**). Overall, the set of four phenotypes was better represented in our study than in previous studies (**Suppl. Table 1**), which stratified subjects less completely or had fewer individuals in total (Tsuang *et al.*).

**Table 1.** Demographics from participants included in the current study, pathologically evaluated in the National Alzheimer’s Coordinating Center (NACC) or Rush University Medical Center databases. . AD^+^LB^+^ corresponds to AD-LB co-pathology individuals, AD^+^LB^−^ corresponds to sole AD pathology individuals, AD^−^LB^+^ corresponds to sole LB pathology individuals, and AD^−^LB^−^ corresponds to individuals with neither pathology. AAD is age-at-death, reported as mean ± standard deviation.

|  | Overall |  |  | NACC |  |  | Rush |  |  |
| --- | --- | --- | --- | --- | --- | --- | --- | --- | --- |
|  | N | N female | AAD | N | N female | AAD | N | N female | AAD |
| **AD+LB+** | 1,072 | 544 (51%) | 82.9 ± 9.3 | 851 | 395 (46%) | 80.8 ± 8.9 | 221 | 149 (67%) | 91.0 ± 5.8 |
| **AD+LB-** | 2,492 | 1,435 (58%) | 83.5 ± 9.4 | 1,973 | 1,055 (53%) | 81.5 ± 9.2 | 519 | 380 (73%) | 91.1 ± 5.7 |
| **AD-LB+** | 158 | 76 (48%) | 86.6 ± 8.1 | 84 | 34 (40%) | 85.4 ± 8.9 | 74 | 42 (57%) | 88.0 ± 6.8 |
| **AD-LB-** | 1,263 | 706 (56%) | 87.3 ± 8.1 | 724 | 376 (52%) | 86.8 ± 8.8 | 539 | 330 (61%) | 87.9 ± 7.1 |

### Genome-wide analysis

We performed a meta-analysis of separate GWAS in the NACC and Rush subsets of our cohort for each of AD^+^LB^+^, AD^+^LB^−^, and AD^−^LB^+^ pathology compared to AD^−^LB^−^ pathology, as well as for AD^+^LB^+^ versus AD^+^LB^−^ pathology. We used PLINK 2.0 for logistic regression and included sex, age-at-death, and the top ten principal components accounting for genetic ancestry as covariates [11]. We removed duplicates and first-degree relatives within and between genomic datasets using KING [37]. In each pair of relatives, the relative with younger age at death was preferentially kept or the oldest control in the absence of pathology cases. We filtered out genetic variants that had a minor allele frequency below 0.01, departed from Hardy-Weinberg equilibrium with a significance below *P <* 10^−5^, or had a missingness rate above 20%. We imputed data on the TOPMed reference panel as described in [33] and considered variants with R^2^ > 0.8. We meta-analyzed the separate NACC and Rush GWAS using the inverse variance weighted method in METAL [55]. Manhattan plots from summary statistics were produced using the R package CMplot [56]. The significance threshold was set at *P <* 5 × 10^−8^, the standard threshold for genome-wide significance. We estimated the association of *APOE*-*ε*4 with risk of AD^+^LB^+^, AD^+^LB^−^, and AD^−^LB^+^ pathology relative to AD^−^LB^−^ pathology, and the association with risk of AD^+^LB^+^ pathology relative to AD^+^LB^−^ pathology, in terms of odds ratio (OR). We also estimated the association of *APOE*-*ε*2. We compared our estimates to those in the literature [52, 28, 13, 2, 10, 14, 18, 46, 48] and when relevant we computed measures of linkage disequilibrium between variants in European ancestry populations using LDlink [35]. We examined loci besides *APOE* that led to genome-wide significant signals. We explored lead variant annotation at significant loci using gnomAD [45]. Finally, we surveyed AD and PD risk loci reported in large clinical case-control GWAS [3, 12, 40] and report the ones associated with pathology at the nominal significance level (*P* < 0.05) in our study.

## Results

We observed that *APOE*-*ε*4 (rs429358) was associated with risk of AD^+^LB^+^ pathology versus AD^−^LB^−^ pathology (OR = 4.24, 95% CI = 3.52–5.10, *P* = 1.5 × 10^−52^) and risk of AD^+^LB^−^ pathology versus AD^−^ LB^−^ pathology (OR = 4.22, 95% CI = 3.60–4.96, *P* = 1.4 × 10^−69^) (**Fig. 2a–b**; **Table 4**). We did not observe an association of *APOE*-*ε*4 with the risk of AD^−^LB^+^ pathology versus AD^−^LB^−^ pathology (OR = 0.93, 95% CI = 0.60–1.43, *P* = 0.73) or risk of AD^+^LB^+^ pathology versus AD^+^LB^−^ pathology (OR = 1.01, 95% CI = 0.90–1.13, *P* = 0.83) (**Fig. 2c–d**; **Table 4**). Another gene locus that yielded significant associations was *BIN1*. Like *APOE*-*ε*4, we observed that rs4663105 on the *BIN1* locus was associated with risk of AD^+^LB^−^ pathology compared to AD^−^LB^−^ pathology (OR = 1.40, 95% CI = 1.26–1.56, *P* = 6.5 × 10^−10^) and risk of AD^+^LB^+^ pathology compared to AD^−^LB^−^ pathology (OR = 1.53, 95% CI = 1.35–1.75, *P* = 1.4 × 10^−10^) (**Fig. 2a–b**; **Table 5**). rs4663105 was not observed to be associated with risk of AD^−^ LB^+^ pathology versus AD^−^LB^−^ pathology (OR = 1.10, 95% CI = 0.85–1.41, *P* = 0.48) or risk of AD^+^LB^+^ pathology versus AD^+^LB^−^ pathology (OR = 1.13, 95% CI = 1.02–1.25, *P* = 0.019) at the genome-wide significance level (**Fig. 2c–d**; **Table 5**). When using pathological criteria from Tsuang *et al.* (2013) (**Fig. 1a**), effect estimates for *APOE*-*ε*4 differed considerably from those reported in the original study, particularly so for the effect on AD^−^LB^+^ vs. AD^−^LB^−^ (**Table 4**). On the contrary, there was fair agreement when using pathological criteria from Kaivola *et al.* (2022) (**Fig. 1b**; **Table 4**). *APOE*-*ε*2 showed similar results to *APOE*-*ε*4, except with the opposite direction of effect in the GWAS where *APOE*-*ε*4 exhibited an association (**Suppl. Table 3**). Overall, we observed an enrichment among the 79 variants listed in the clinical AD GWAS (Bellenguez *et al.* (2022)) and tested in our analyses; we observed an enrichment of nominally significant associations with concordant direction of effect: 20.3% variants (16/79) in the AD^+^LB^−^ vs. AD^−^LB^−^ contrast and 24.1% (19/79) in the AD^+^LB^+^ vs. AD^−^ LB^−^ (with the chance level being at 2.5%). In contrast, we did not observe a significant enrichment for the 76 variants identified in PD clinical GWAS Chang *et al.* (2017) and Nalls *et al.* (2019): 2.6% (2/76) in the AD^+^LB^+^ vs. AD^−^LB^−^ contrast, 3.9% (3/76) in the AD^−^LB^+^ vs. AD^−^LB^−^ contrast, and 2.6% (2/76) in the AD^+^LB^+^ vs. AD^+^LB^−^ contrast. Among known AD risk loci besides *BIN1* and *APOE* reported by Bellenguez *et al.* (2022), *ADAM17* (rs72777026), *COX7C* (rs62374257), *HLA* (rs6605556), *TREM2* (rs143332484), *HS3ST5* (rs785129), *SEC61G* (rs76928645), *CLU* (rs11787077), *ECHDC3* (rs7912495), *TPCN1* (rs6489896), *FERMT2* (rs17125924), *DOC2A* (rs1140239), *PRDM7* (rs56407236), *ABI3* (rs616338), *ABCA7* (rs12151021), and *SIGLEC11* (rs9304690) were concordant and nominally associated with AD^+^LB^+^ versus AD^−^LB^−^; and *CR1* (rs679515), *ADAM17* (rs72777026), *INPP5D* (rs10933431), *CLNK/HS3ST1* (rs6846529), *ANKH* (rs112403360), *COX7C* (rs62374257), *HLA* (rs6605556), *TREM2* (rs143332484), *ZCWPW1/NYAP1* (rs7384878), *PTK2B* (rs73223431), *CLU* (rs11787077), *ECHDC3* (rs7912495), *PICALM* (rs3851179), *SORL1* (rs11218343), *FERMT2* (rs17125924), *APH1B* (rs117618017), *MAF* (rs450674), and *ABCA7* (rs12151021) were concordant and nominally associated with AD^+^LB^−^ pathology versus AD^−^LB^−^ (**Table 6; Suppl. Table 5**). Among known PD risk loci reported by Chang *et al.* (2017) and Nalls *et al.* (2019), *SCN3A* (rs353116) and *HLA-DRB6/HLA-DQA1* (rs9275326) were concordant and nominally associated with AD^+^LB^+^ versus AD^−^LB^−^; *TMEM175/DGKQ* (rs34311866), *FAM200B/CD38* (rs11724635), and *SNCA* (rs356182) were concordant and nominally associated with AD^−^LB^+^ versus AD^−^LB^−^; and *GBA* (rs35749011) and *TMEM175/DGKQ* (rs34311866) were concordant and nominally associated with AD^+^LB^+^ versus AD^+^LB^−^ (**Table 6; Suppl. Table 5**). Notably, the *TPCN1* locus, reported to be associated with LB dementia by Kaivola *et al.* (2023), was associated with AD^+^LB^+^ pathology and AD^+^LB^−^ pathology versus AD^−^LB^−^ pathology below or near the nominal significance level, but not so with AD^−^LB^+^ pathology versus AD^−^LB^−^ pathology or AD^+^LB^+^ pathology versus AD^+^LB^−^ pathology.

**Figure 2.**
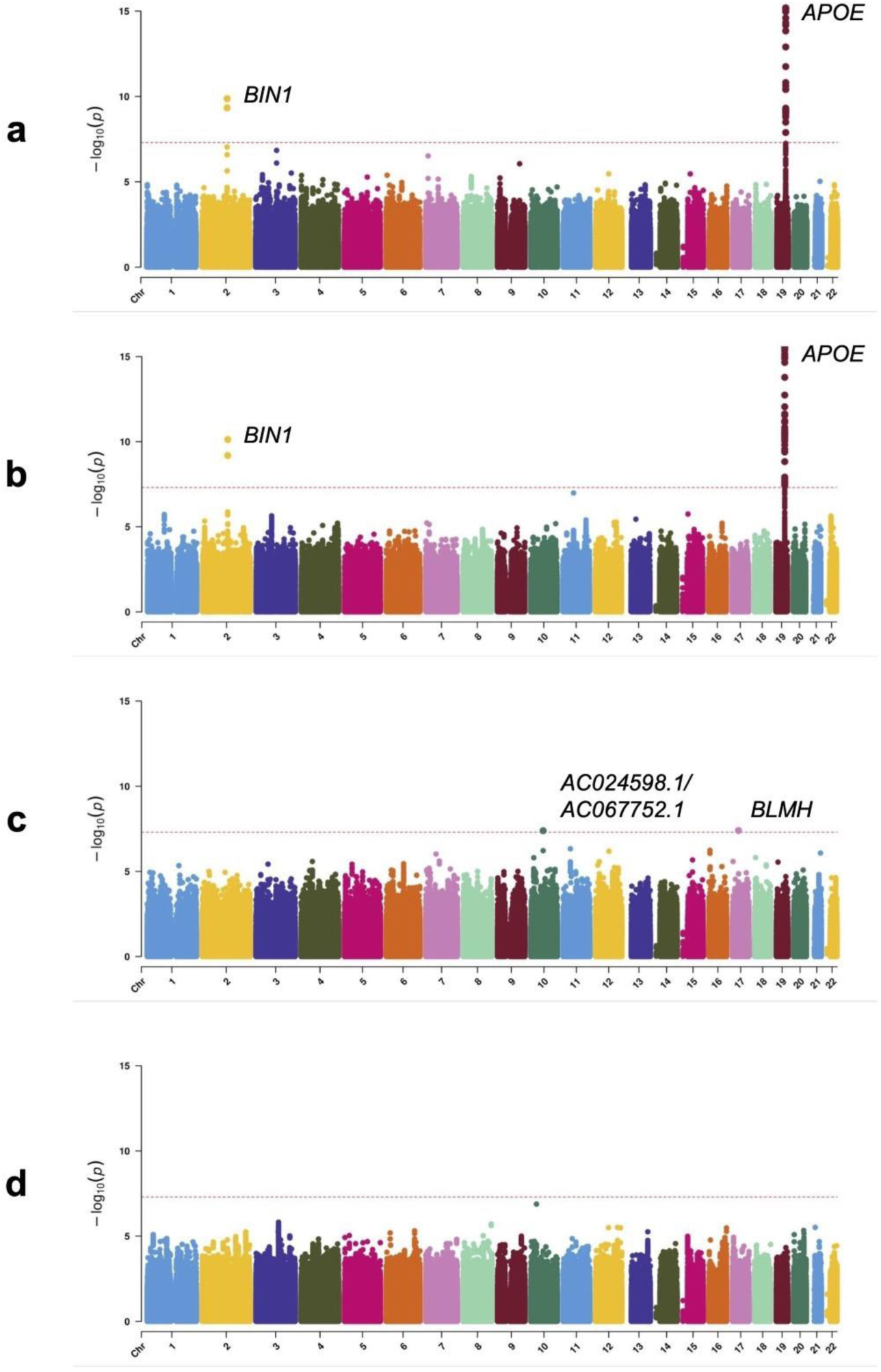
Manhattan plots of genetic association with pathology contrasts. **a.** Association with AD^+^LB^+^ pathology versus AD^−^LB^−^ pathology. **b.** Association with AD^+^LB^−^ pathology versus AD^−^LB^−^ pathology. **c.** Association with AD^−^LB^+^ pathology versus AD^−^LB^−^ pathology. **d.** Association with AD^+^LB^+^ pathology versus AD^+^LB^−^ pathology. Variants at two novel loci exhibited genome-wide significant associations in the AD^−^LB^+^ versus AD^−^LB^−^ analysis (rs112017605 on both an intron of *AC024598.1* and an intron of *AC067752.1* on chromosome 10 and rs116691607 on an intron of *BLMH* on chromosome 17) (**c**; **Suppl. Table 2**), but we do not discuss these candidates in the main text because neither was flanked by a set of nearby variants in linkage disequilibrium, raising concern that these could be spurious signals.

**Table 2.** *APOE*-*ε*4 allele frequency by pathology category. The second and third columns indicate the frequency of *APOE-ε4* among the NACC or Rush individuals in each category. Frequencies are reported as count of *APOE-ε4* alleles out of total allele count.

|  | Overall | NACC | Rush |
| --- | --- | --- | --- |
| **AD+LB+** | 744/2,144 (34.7%) | 662/1,702 (38.9%) | 82/442 (18.6%) |
| **AD+LB-** | 1,685/4,984 (33.8%) | 1,492/3,946 (37.8%) | 193/1,038 (18.6%) |
| **AD-LB+** | 28/316 (8.9%) | 20/168 (11.9%) | 8/148 (5.4%) |
| **AD-LB-** | 249/2,526 (9.9%) | 166/1,448 (11.5%) | 83/1,078 (7.7%) |

**Table 3.** Number of individuals in pathology categories across analyses. Not all classifications were necessarily pathologically confirmed (**Suppl.** Table 1). First column: our study cohort. Next two columns: our preliminary cohort (before the removal of individuals not classified by our criteria) classified using literature criteria (**Fig. 1a–b**) [52, 28]. Remaining columns: category sizes in literature cohorts [52, 28, 13, 2, 10, 14, 18, 24, 46, 48]. For Beecham *et al.,* Robinson *et al.,* and Sabir *et al.,* only analyses of *APOE*-*ε*4-associated risk for LB pathology or dementia are considered [2, 46, 48]. For Guerreiro *et al.,* we describe the larger discovery cohort [24].

|  | Current sample | Initial sample × Tsuang *et al.* (2013) criteria | Initial sample × Kaivola *et al.* (2022) criteria | Tsuang *et al.* (2013) | Kaivola *et al.* (2022) | Chia *et al.* (2021) |
| --- | --- | --- | --- | --- | --- | --- |
| AD+LB+ | 1,072 | 916 | 695 | 224 | 66/341<sup>a</sup> | ? |
| AD+LB- | 2,492 | 2,492 | 3,493 | 244 | 0 | 0 |
| AD-LB+ | 158 | 316 | 158 | 91 | 88 | ? |
| AD-LB- | 1,263 | 1,358 | 908 | 269 | 2,928 | 4,027 |
| LB+<sup>b</sup> | N/A | N/A | N/A | N/A | N/A | 2,591 |
| LB-<sup>b</sup> | N/A | N/A | N/A | N/A | N/A | N/A |

|  | Beecham *et al.* (2014) | Bras *et al.* (2014) | Chung *et al.* (2015) | Dickson *et al.* (2018) | Guerreiro *et al.* (2018) | Robinson *et al.* (2018) | Sabir *et al.* (2019) |
| --- | --- | --- | --- | --- | --- | --- | --- |
| AD+LB+ | ? | ? | 215 | 10/27/115/19/<br>111/209<sup>a</sup> | ? | 130/96<sup>a,c</sup> | ? |
| AD+LB- | ? | 0 | 316 | 0 | 0 | 16/60<sup>a</sup> | 0 |
| AD-LB+ | ? | ? | 0 | 46/80/33<sup>a</sup> | ? | 10/12/22<sup>a</sup> | ? |
| AD-LB- | ? | 2,624 | 0 | 660 | 3,791 | 0 | 591 |
| LB+<sup>b</sup> | 2,391 | 667 | N/A | N/A | 1,216 | N/A | 525 |
| LB-<sup>b</sup> | 1,135 | N/A | N/A | N/A | N/A | N/A | N/A |

<sup>a</sup>In Dickson *et al.*, the AD+LB+ individuals were subdivided into individuals with moderate or high AD pathology and brainstem, transitional, or diffuse LB pathology; the subgroup sizes are listed in the order moderate-brainstem, moderate-transitional, *et cetera* [18]. In Robinson *et al.*, the AD+LB+ individuals were subdivided into individuals with primary intermediate or high AD pathology and secondary LB pathology and individuals with primary brainstem, limbic, or neocortical LB pathology [46]. In Kaivola *et al.*, the AD+LB+ individuals were subdivided into individuals with intermediate or high AD pathology [28]. A separate analysis was performed on each subgroup in each of these three studies. <sup>b</sup>These rows are populated only if an analysis was performed on an LB- or LB+ group. In this case the sizes of the corresponding subgroups are
marked as unknown (*e.g.*, AD+LB<sup>+</sup> and AD-LB<sup>+</sup> if LB<sup>+</sup> is known). <sup>c</sup>The six subgroups of this category were consolidated into two subgroups analyzed separately for association of *APOE-ε4* with AD co-pathology versus sole intermediate or high AD pathology or LB co-pathology versus sole brainstem, limbic, or neocortical LB pathology: 130 individuals with primary AD pathology and secondary LB pathology and 96 with primary LB pathology and secondary AD pathology, respectively [46].

**Table 4.**
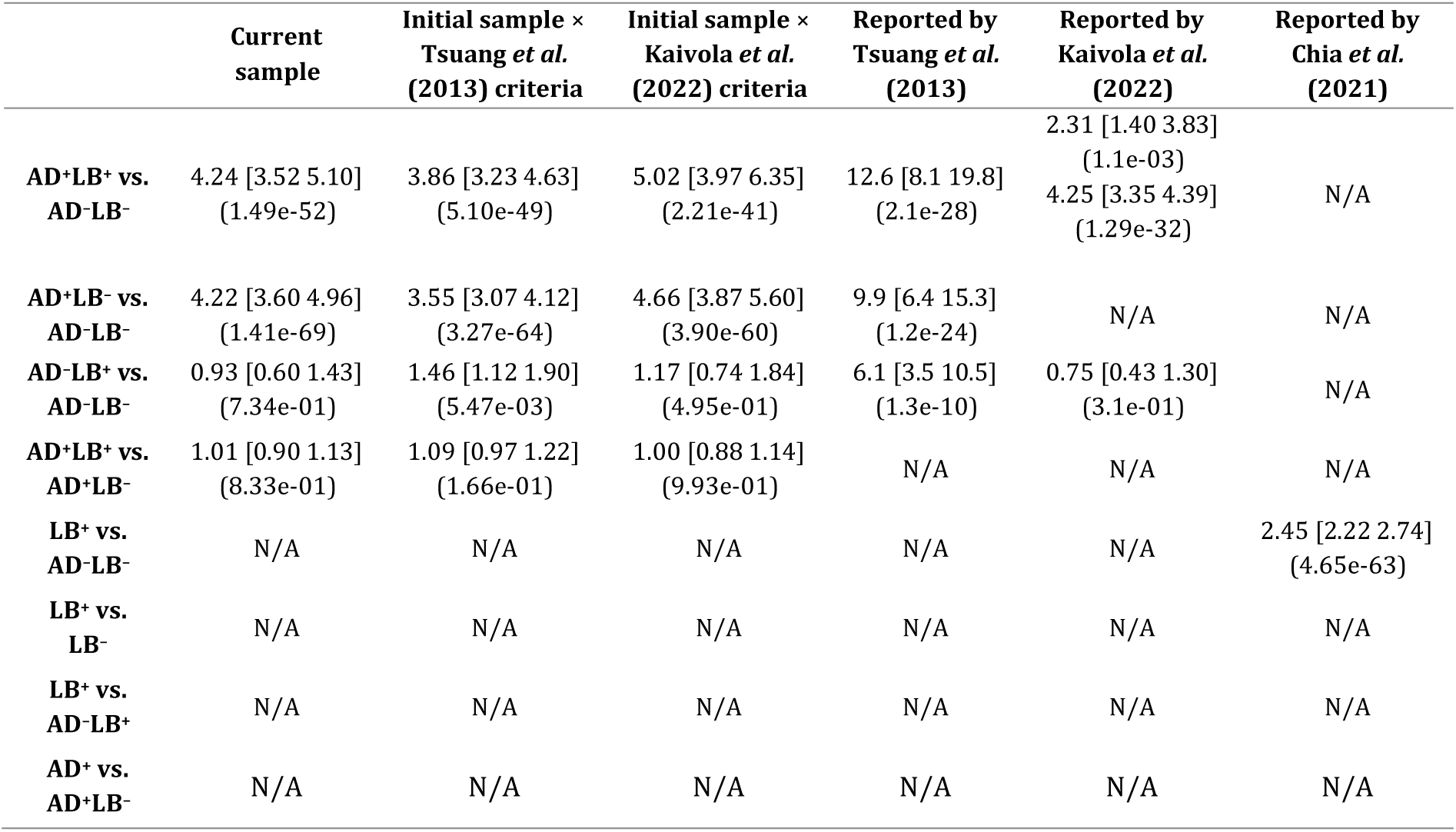
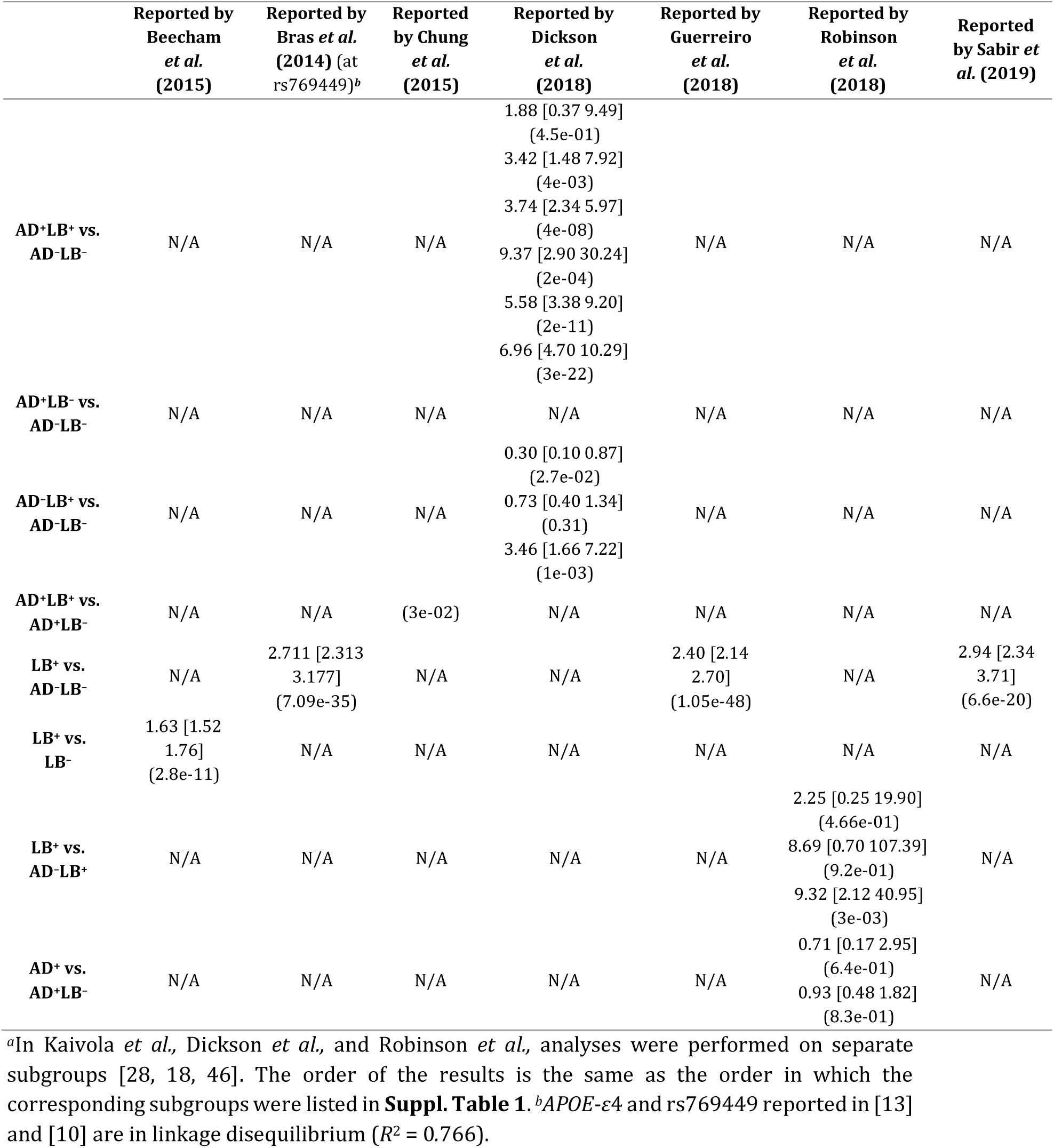
Association of *APOE*-*ε*4 (rs429358) with different pathology contrasts. The first column corresponds to the current study, while the following two columns correspond to results obtained using the current sample using literature criteria to classify participants into pathology groups (**Fig. 1a–b**) [52, 28]. The other columns correspond to results reported in the literature [52, 28, 13, 2, 10, 14, 18, 24, 46, 48]. Effect sizes are reported as OR with 95% confidence interval [CI] and significance (*P*-value).

**Table 5.** Association of rs4663105 on the *BIN1* locus with different pathology contrasts. The first column corresponds to the current study, while the following two columns correspond to results obtained using the current sample using literature criteria to classify participants into pathology groups (**Fig. 1a–b**) [52, 28]. The last column corresponds to a result reported in the literature [13]. Effect sizes are reported as OR with 95% confidence interval [CI] and significance (*P*-value).

|  | Current study | Initial sample ×<br>Tsuang *et al.*<br>(2013) criteria | Initial sample ×<br>Kaivola *et al.*<br>(2022) criteria | Reported by Chia<br>*et al.* (2021)<br>(at rs6733839)<sup>a</sup> |
| --- | --- | --- | --- | --- |
| **AD+LB<sup>+</sup> vs.**<br>**AD-LB<sup>-</sup>** | 1.53 [1.35 1.75]<br>(1.35e-10) | 1.55 [1.35 1.77]<br>(2.19e-10) | 1.56 [1.33 1.82]<br>(1.99e-08) | N/A |
| **AD+LB<sup>-</sup> vs.**<br>**AD-LB<sup>-</sup>** | 1.40 [1.26 1.56]<br>(6.51e-10) | 1.36 [1.23 1.51]<br>(4.45e-09) | 1.36 [1.22 1.52]<br>(6.32e-08) | N/A |
| **AD-LB<sup>+</sup> vs.**<br>**AD-LB<sup>-</sup>** | 1.10 [0.85 1.41]<br>(4.76e-01) | 1.20 [1.00 1.44]<br>(4.81e-02) | 1.11 [0.86 1.43]<br>(4.18e-01) | N/A |
| **AD+LB<sup>+</sup> vs.**<br>**AD+LB<sup>-</sup>** | 1.13 [1.02 1.25]<br>(1.91e-02) | 1.14 [1.03 1.28]<br>(1.57e-02) | 1.20 [1.07 1.35]<br>(2.10e-03) | N/A |
| **LB<sup>+</sup> vs.**<br>**AD-LB<sup>-</sup>** | N/A | N/A | N/A | 1.25 [1.16 1.35]<br>(4.16e-09) |

<sup>a</sup>rs4663105 and rs6733839 reported in [13] are in linkage disequilibrium ( $R^2 = 0.8968$ ).

**Table 6.**
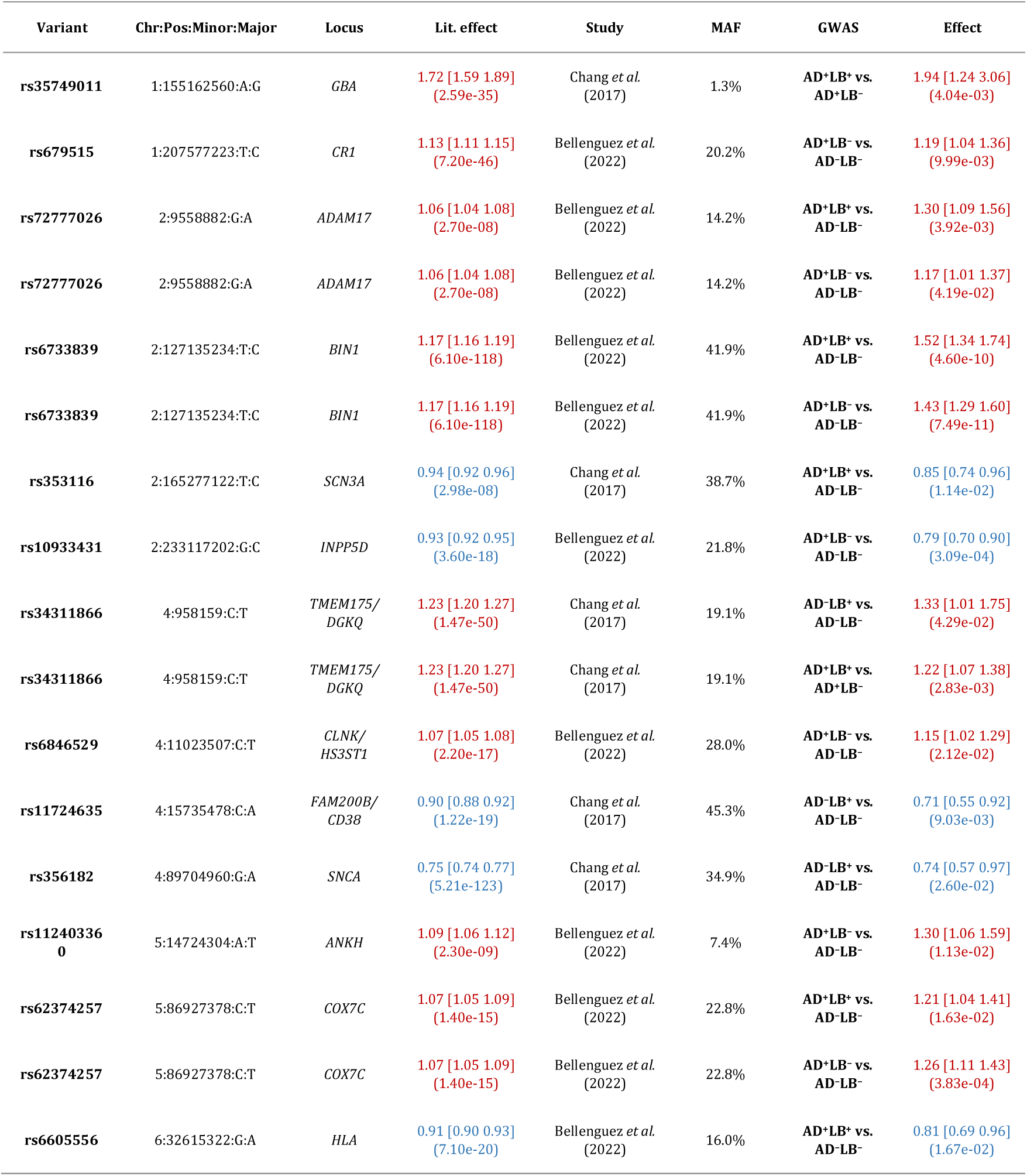
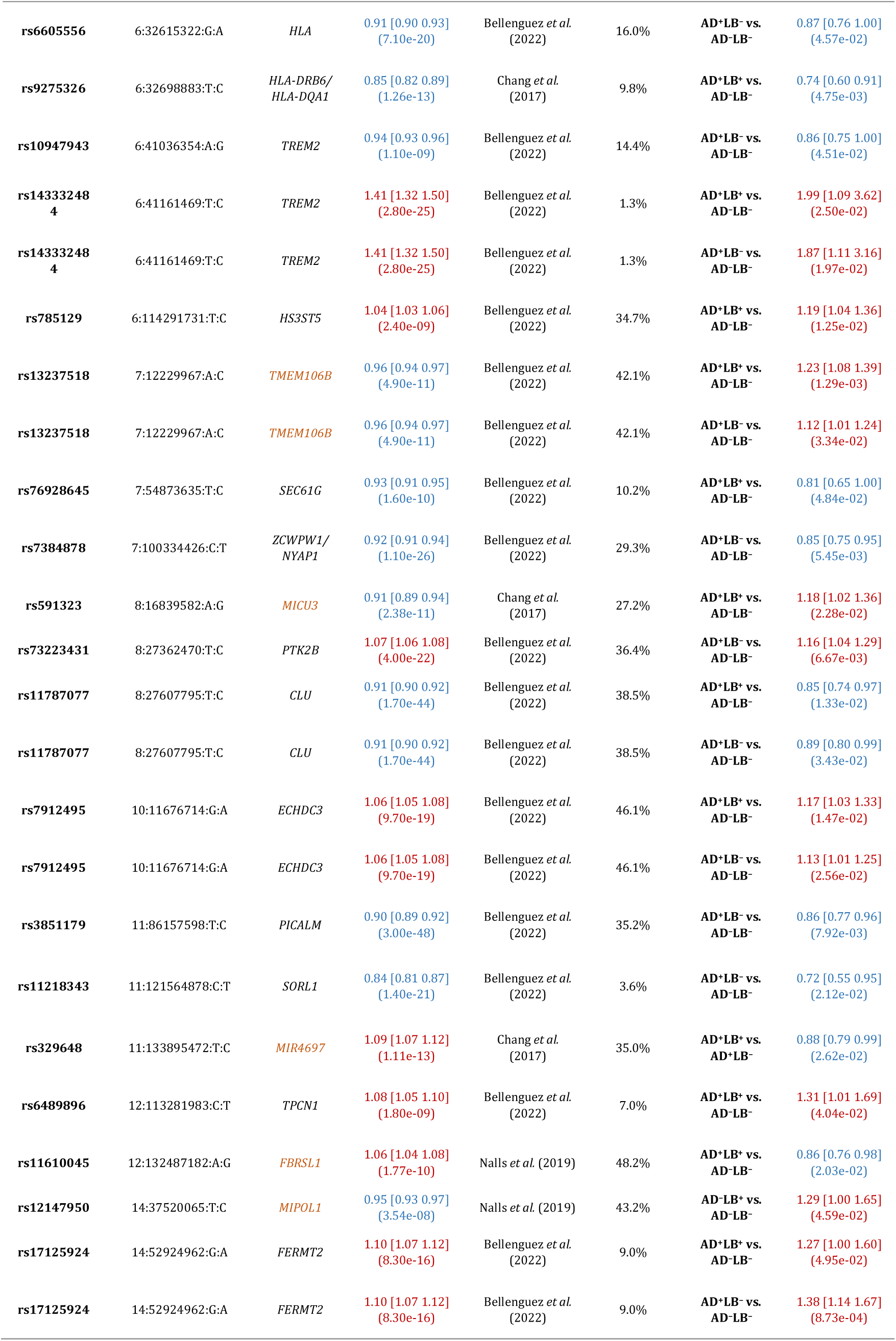
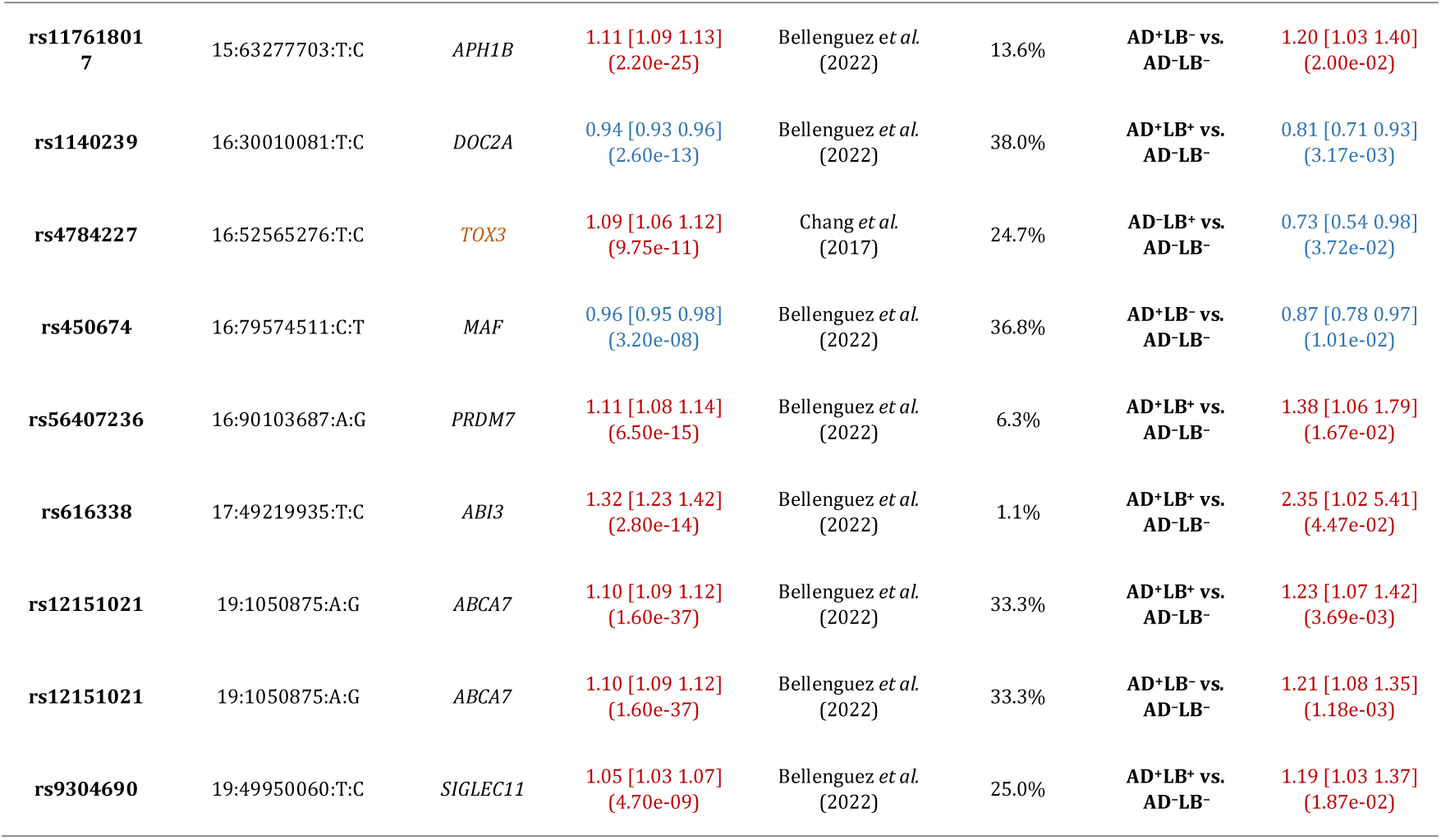
Known Alzheimer’s disease risk loci reported in Bellenguez *et al.* (2022) and known Parkinson’s disease risk loci reported in Chang *et al.* (2017) and Nalls *et al.* (2019) which are associated with the corresponding pathology contrasts at the nominal significance level (*P* < 0.05) [3, 12, 40]. For AD risk loci, associations with AD^+^LB^+^ vs. AD^−^LB^−^ or AD^+^LB^−^ vs. AD^−^LB^−^ are shown. For PD risk loci, associations with AD^+^LB^+^ vs. AD^−^LB^−^, AD^−^LB^+^ vs. AD^−^LB^−^, or AD^+^LB^+^ vs. AD^+^LB^−^ are shown. Chr:Pos:Minor:Major is chromosome, position (genome build hg38, GRCh38), and minor allele and major allele in our study. Lit. effect is the effect size reported in the literature. MAF is the minor allele frequency in our study. Loci with results discordant in terms of direction of effect are colored orange.

## Discussion

Our study emphasizes that *APOE*-*ε*4 is associated with the risk of both AD^+^LB^−^ pathology (OR = 4.22, *P* = 1.4 × 10^−69^) and AD^+^LB^+^ co-pathology (OR = 4.24, *P* = 1.5 × 10^−52^) compared to AD^−^LB^−^ pathology. These OR estimates were lower than the estimates in Tsuang *et al.* for *APOE*-*ε*4-associated risk of AD^+^LB^−^ pathology (OR = 12.6, *P* = 2.1 × 10^−28^) and AD^+^LB^+^ co-pathology (OR = 9.9, *P* = 1.2 × 10^−24^) (**Table 4**) [52]. Tsuang *et al.* appear to have overestimated the true effect size of *APOE*-*ε*4 in their positive pathology subjects, possibly due to the smaller size of their cohort [52] (N = 640 versus N = 4,985 in the current study) (**Table 3**). Our estimate of *APOE*-*ε*4-associated risk for AD^+^LB^+^ pathology is in line with the estimate in Kaivola *et al.,* the next largest study of pathologically assessed individuals (OR = 4.25, *P* = 1.29 × 10^−32^) [28]. Our data substantiate that *APOE*-*ε*4 is a driver of AD pathology. Notably, we estimated a similar effect size of *APOE*-*ε*4 on the risk of AD^+^LB^−^ pathology (OR = 4.22) and on the risk of AD^+^LB^+^ co-pathology (OR = 4.24) versus AD^−^LB^−^ pathology, suggesting that *APOE*-*ε*4 does not have a specific effect on the development of LB pathology in individuals with AD pathology. Consistent with this finding, we did not observe *APOE*-*ε*4 to be associated with the risk of AD^+^LB^+^ co-pathology compared to sole AD^+^LB^−^ pathology (OR = 1.01, *P* = 0.83).

This finding contradicts the results presented by Chung *et al.* (2015), where *APOE*-*ε*4 was found to be associated with AD^+^LB^+^ co-pathology when compared to sole AD^+^LB^−^ pathology (*P* = 0.03) [14]. However, their AD^+^LB^+^ group was five times smaller than ours (N = 215 versus N = 1,072), and the AD^+^LB^−^ group was eight times smaller (N = 316 versus N = 2,492). Our result is consistent with the finding in Robinson *et al.* (2018) that *APOE*-*ε*4 is not associated with the co-occurrence of AD pathology with other pathologies (OR = 0.71, *P* = 0.64 for intermediate AD pathology; and OR = 0.93, *P* = 0.83 for high AD pathology) [46]. Our result is also consistent with the finding in Dickson *et al.* (2018) that *APOE*-*ε*4 is not associated with higher Lewy body count in individuals with moderate AD pathology (*P* ≥ 0.30 for all regions) or high AD pathology (*P* ≥ 0.069 for all regions) [18]. Taken together, *APOE*-*ε*4 appears similarly prevalent in AD pathology cases with or without LB pathology. Furthermore, we did not find *APOE*-*ε*4 to be associated with risk for sole LB pathology (AD^−^LB^+^) pathology (OR = 0.93, *P* = 0.73) compared to no pathology (AD^−^LB^−^). This is in contradiction with [52] where *APOE*-*ε*4 was strongly associated with risk for AD^−^LB^+^ pathology (OR = 6.1, *P* = 1.3 × 10^−10^). This discrepancy could be because Tsuang *et al.* used a more stringent definition of AD pathology (**Fig. 1a**), leaving open the possibility that individuals whom we would have classified as AD^+^LB^+^ with our criteria were instead classified as AD^−^LB^+^. Indeed, when we categorized our initial pathologically evaluated cohort using the exact same criteria as in [52], testing the association of *APOE*-*ε*4 with risk of AD^−^LB^+^ pathology yielded a modestly higher OR and nominal significance (OR = 1.46, *P* = 5.5 × 10^−3^) (**Table 4**). The main difference between our criteria and those of Tsuang *et al.* is that we lower the threshold for AD pathology to Braak stage III NFTs plus sparse neuritic plaques in LB^+^ individuals, suggesting that LB pathology commonly occurs in *APOE*-*ε*4 carriers with potential early-stage AD but not in firmly non-AD *APOE*-*ε*4 carriers. Knowing the breakdown of the *APOE*-*ε*4-positive subjects in [52] by Braak stage and CERAD score would establish further support for this interpretation. Dickson *et al.* also found an association of *APOE*-*ε*4 with the risk of diffuse LB pathology and low AD pathology (OR = 3.46, *P* = 0.001) while classifying individuals with Braak stage III NFTs and Thal phase 0, 1, or 2 amyloid-β non-neuritic plaques as having low AD pathology [18]. In this AD^−^LB^+^ group, the median Braak stage was III and the median Thal phase was 1. We would have classified this subset as AD^+^LB^+^. A Thal phase of 1 tends to correspond to a CERAD score of sparse neuritic plaques or higher [7]. Therefore, many individuals in this AD^−^LB^+^ group in [18] had a Braak stage of III and at least sparse neuritic plaques; this subset was likely the source of the elevated frequency of *APOE*-*ε*4 in the group. We would have classified this subset as AD^+^LB^+^ instead. It should be noted that this AD^−^LB^+^ group in Dickson *et al.* was smaller than ours (N = 54 versus N = 158) and the controls were not pathologically confirmed. Another cause of the discrepancy between our result that *APOE*-*ε*4 was not associated with the risk of AD^−^LB^+^ pathology and Tsuang *et al.*’s finding that *APOE*-*ε*4 increased risk for LB pathology may have been that the pathologically confirmed AD^−^LB^−^ group in [52] was more than four times smaller than ours (N = 269 versus N = 1,263). Remarkably, the frequency of *APOE*-*ε*4 was 31.9% in the AD^−^LB^+^ group [52], which was far higher than in our AD^−^LB^+^ group (8.9%). Our result was consistent with the finding in Dickson *et al.* that *APOE*-*ε*4 was not associated with the risk of the AD^−^LB^+^ phenotype of transitional (limbic) LB pathology and low AD pathology (OR = 0.73, *P* = 0.31) [18].

In a larger study than [52] of pathologically confirmed LB dementia cases and mixed pathologic and clinical controls, Chia *et al.* found that *APOE*-*ε*4 was associated with risk of LB dementia: OR = 2.45 with *P* = 4.65 × 10^−63^ for rs769449, which is in linkage disequilibrium with *APOE*-*ε*4 with *R*^2^ = 0.766 [13]. However, this could have been because Chia *et al.* did not categorize individuals by AD pathology status, and many AD^+^LB^+^ individuals were inevitably included in the LB^+^ case group. When Kaivola *et al.* studied the cohort in Chia *et al.* using more precise pathological categorization, *APOE*-*ε*4 was not observed to have a significant effect on risk for AD^−^LB^+^ pathology (OR = 0.75, *P* = 0.31) [28]. Associations of *APOE*-*ε*4 with risk of LB pathology (OR = 1.63, *P* = 2.8 × 10^−11^) and LB dementia (OR = 2.71, *P* = 7.1 × 10^−35^; OR = 2.40, *P* = 1.05 × 10^−48^; and OR = 2.94, *P* = 6.6 × 10^−20^) reported in Beecham *et al.* (2014), Bras *et al.* (2014), Guerreiro *et al.* (2018), and Sabir *et al.* (2019), respectively, could similarly have been because these studies did not exclude AD^+^LB^+^ individuals from their LB^+^ case groups [2, 10, 24, 48]. Our finding was also consistent with the result in Robinson *et al.* that *APOE*-*ε*4 was associated with the co-occurrence of cortical LB pathology with other pathologies, including AD pathology, versus sole cortical LB pathology (OR = 9.32, *P* = 0.003) [46]. The latter result would imply that *APOE*-*ε*4 was rarer in the AD^−^LB^+^ individuals in [46] than in LB^+^ individuals with advanced LB pathology. Presumably, because 80% of LB^+^ individuals also had AD pathology, the prevalence of *APOE*-*ε*4 in LB^+^ individuals was most likely unrelated to the presence of LB pathology.

The balance of evidence thus suggests that *APOE*-*ε*4 does not affect risk for AD^−^LB^+^ pathology when strictly defined to exclude possible early-stage AD. Subjects with LB pathology and no AD pathology have been rare, and more are needed to substantiate this conclusion. This concept does not oppose the observations in Dickson *et al.* and Zhao *et al.* (2020) that *APOE*-*ε*4 was associated with higher LB counts in AD^−^LB^+^ subjects, as neither analysis compared cases to controls without pathology [18, 57]. Similarly compatible is the observation in Goldberg *et al.* (2020) that *APOE*-*ε*4 was associated with further propagated Lewy bodies; although Goldberg *et al.* adjusted for AD pathology level, their analysis did not specifically compare LB^+^ to LB^−^ subjects [23]. It is conceivable that *APOE*-*ε*4 worsens LB pathology but does not influence its actual emergence in individuals without AD pathology. Overall, our data suggest *APOE*-*ε*4 is most likely not involved in the emergence of LB pathology in the absence or presence of AD pathology. This interpretation is further supported by the lack of an effect of *APOE*-*ε*4 in the largest GWAS of clinically defined PD (OR = 1.02, *P* = 0.49) [40].

A second gene locus that yielded significant associations was *BIN1.* As for *APOE*-*ε*4, the *BIN1* lead variant was associated with the risk of sole AD (AD^+^LB^−^) and AD^+^LB^+^ co-pathology, but not sole LB (AD^−^LB^+^) when compared to no pathology (AD^−^LB^−^). *BIN1* was also not associated with the risk of AD^+^LB^+^ co-pathology when compared to sole AD pathology (AD^+^LB^−^) pathology. These results further corroborate that *BIN1* is also a driver of AD pathology. In the largest previous GWAS of LB pathology, Chia *et al.* found that *BIN1* is a risk locus for pathologically confirmed LB dementia (OR = 1.25, *P* = 4.16 × 10^−9^ for rs6733839, in linkage disequilibrium with rs4663105 with *R*^2^ = 0.8968) [13]. However, when gathering individuals, Chia *et al.* did not select against AD pathology, which was presumably far more prevalent in the LB^+^ case group than in the control group. Given that we do not observe an association of rs4663105 with risk for AD^−^LB^+^ pathology, the *BIN1* association reported in [13] may have been driven by the AD^+^LB^+^ subgroup within the LB^+^ group. However, our findings are limited by the size of our AD^−^LB^+^ group (N = 158); the lower statistical power of the AD^−^LB^+^ pathology versus AD^−^LB^−^ pathology GWAS was likely also the reason that known synucleinopathy risk loci like *GBA* and *SNCA* did not yield genome-wide significant associations in this analysis. Altogether, the current balance of evidence suggests that variants on the *BIN1* locus behave like *APOE*-*ε*4: pathogenic *BIN1* variants increase the overall risk of LB pathology simply by increasing the risk of AD pathology (which is frequently accompanied by LB pathology), but they do not affect the risk of AD^−^LB^+^ pathology or the risk of co-pathology (AD^+^LB^+^) among AD^+^ individuals. It is worth mentioning that the effect of *BIN1* on the risk of AD pathology may be lifestyle-dependent, as we did not observe any association of *BIN1* with pathology in the subset of Rush individuals alone (**Suppl. Fig. 2; Suppl. Table 4**). The monastic life of these subjects likely militates against disease.

Future studies should continue the effort of determining the risk loci for AD pathology, LB pathology, or AD-LB co-pathology using pathologically well-categorized and clinically unbiased cohorts. It may be worth focusing on comparing AD^+^LB^+^ to AD^+^LB^−^ groups to identify LB pathology risk loci because the sample size of either phenotype is larger than AD^−^LB^+^. Further study of the AD^+^LB^+^ versus AD^+^LB^−^ and AD^−^LB^+^ versus AD^−^LB^−^ contrasts may also reveal possible differences between genetic risk factors underlying LB pathology in the presence or absence of AD pathology; we propose a hypothetical genetic model in **Suppl. Fig. 3**.

## Conclusion

In conclusion, our set of GWAS meta-analyses indicates that while *APOE*-*ε*4 is a risk factor for AD pathology and increases risk of AD-LB co-pathology, it is not a risk factor for LB pathology independent of AD pathology or along with AD pathology. This is also true of variants on the *BIN1* locus; therefore, neither *APOE-ε*4 nor *BIN1* variants appear to play a specific mechanistic role in the emergence of LB pathology. We provide GWAS meta-analysis summary statistics that will enable more reliable, pathologically precise polygenic risk score calculations for AD, LB dementia, and related disorders. Ultimately, we shed light on the genetic bases of AD and LB pathology, which will be useful for further unraveling the etiology of these debilitating pathologies and developing accurate and effective interventions.

## Acknowledgments

This research is based on data collected by the National Alzheimer’s Coordinating Center and the Rush University Medical Center.

## Availability of data and materials

The summary statistics from our genome-wide association study meta-analyses are available online (URL to NIAGADS and EBI GWAS catalogue to be updated at publication).

## Authors’ contributions

Concept and design: YLG, MDG. Data analysis: ST and YLG. Drafting of the manuscript: ST, YLG, NK, MDG. Critical review of the manuscript: ST, YLG, NK, MEB, and MDG. Funding acquisition: YLG, MEB, and MDG.

## Declarations

### Ethical approval and consent to participate

Participants or their caregivers provided written informed consent in the original studies. The current study protocol was granted an exemption by the Stanford University institutional review board because the analyses were carried out on deidentified data; therefore, additional informed consent was not required.

## References

1. Beach TG, Monsell SE, Phillips LE, Kukull W (2012) Accuracy of the Clinical Diagnosis of Alzheimer Disease at National Institute on Aging Alzheimer Disease Centers, 2005–2010. J Neuropathol Exp Neurol 71:266–273. https://doi.org/10.1097/NEN.0b013e31824b211b

2. Beecham GW, Hamilton K, Naj AC, Martin ER, Huentelman M, Myers AJ et al (2014) Genome-wide association meta-analysis of neuropathologic features of Alzheimer’s disease and related dementias. PLoS Genet 10. https://doi.org/10.1371/journal.pgen.1004606

3. Bellenguez C, Küçükali F, Jansen IE et al (2022) New insights into the genetic etiology of Alzheimer’s disease and related dementias. Nat Genet 54:412–436. https://doi.org/10.1038/s41588-022-01024-z

4. Belloy ME, Napolioni V, Greicius MD (2019) A Quarter Century of APOE and Alzheimer’s Disease: Progress to Date and the Path Forward. Neuron 101:820–838. https://doi.org/10.1016/j.neuron.2019.01.056

5. Berge G, Sandro SB, Ronge A et al (2014) Apolipoprotein E ε2 genotype delays onset of dementia with Lewy bodies in a Norwegian cohort. J Neurol Neurosurg Psychiatry 85:1227– 1231. https://doi.org/10.1136/jnnp-2013-307228

6. Blennow K, Zetterberg H (2018) Biomarkers for Alzheimer’s disease: current status and prospects for the future. J Intern Med 284:643–663. https://doi.org/10.1111/joim.12816

7. Boluda S, Toledo JB, Irwin DJ et al (2014) A comparison of Aβ amyloid pathology staging systems and correlation with clinical diagnosis. Acta Neuropathol 128:543–550. https://doi.org/10.1007/s00401-014-1308-9

8. Braak H, Alafuzoff A, Arzberger T, Kretzschmar H, Del Tredici K (2006) Staging of Alzheimer disease-associated neurofibrillary pathology using paraffin sections and immunocytochemistry. Acta Neuropathol 112:389–404. https://doi.org/10.1007/s00401-006-0127-z

9. Braak H, Braak E (1997) Frequency of stages of Alzheimer-related lesions in different age categories. Neurobiol Aging 18:351–357. https://doi.org/10.1016/s0197-4580(97)00056-0

10. Bras J, Guerreiro R, Darwent L, Parkkinen L, Ansorge O et al (2014) Genetic analysis implicates APOE, SNCA and suggests lysosomal dysfunction in the etiology of dementia with Lewy bodies. Hum Mol Genet 23:6139–6146. https://doi.org/10.1093/hmg/ddu334

11. Chang CC, Chow CC, Tellier LCAM, Vattikuti S, Purcell SM, Lee JJ et al (2015) Second-generation PLINK: rising to the challenge of larger and richer datasets. GigaScience 4. https://doi.org/10.1186/s13742-015-0047-8

12. Chang D, Nalls M, Hallgrímsdóttir I et al (2017) A meta-analysis of genome-wide association studies identifies 17 new Parkinson’s disease risk loci. Nat Genet 49:1511–1516. https://doi.org/10.1038/ng.3955

13. Chia R, Sabir MS, Bandres-Ciga S, Saez-Atienzar S, Reynolds RH et al (2021) Genome sequencing analysis identifies new loci associated with Lewy body dementia and provides insights into its genetic architecture. Nat Genet 53:294–303. https://doi.org/10.1038/s41588-021-00785-3

14. Chung EJ, Babulal GM, Monsell SE, Cairns NJ, Roe CM, Morris JC et al (2015) Clinical Features of Alzheimer Disease With and Without Lewy Bodies. JAMA Neurol 72:789–796. https://doi.org/10.1001/jamaneurol.2015.0606

15. Corneveaux JJ, Myers AJ, Allen AN, Pruzin JJ, Ramirez M et al (2010) Association of CR1, CLU and PICALM with Alzheimer’s disease in a cohort of clinically characterized and neuropathologically verified individuals. Hum Mol Genet 19:3295–3301. https://doi.org/10.1093/hmg/ddq221

16. Davis AA et al (2020) APOE genotype regulates pathology and disease progression in synucleinopathy. Sci Transl Med 12. https://doi.org/10.1126/scitranslmed.aay3069

17. DeTure MA, Dickson DW (2019) The neuropathological diagnosis of Alzheimer’s disease. Mol Neurodegeneration 14. https://doi.org/10.1186/s13024-019-0333-5

18. Dickson DW, Heckman MG, Murray ME, Soto AI, Walton RL et al (2018) APOE ε4 is associated with severity of Lewy body pathology independent of Alzheimer pathology. Neurology 91:1182–1195. https://doi.org/10.1212%2FWNL.0000000000006212

19. Escott-Price V, Hardy J (2022) Genome-wide association studies for Alzheimer’s disease: bigger is not always better. Brain Commun 4:125. https://doi.org/10.1093/braincomms/fcac125

20. Escott-Price V, Myers AJ, Huentelman M, Hardy J (2017) Polygenic Risk Score Analysis of Pathologically Confirmed Alzheimer Disease. Ann Neurol 82:311–314. https://doi.org/10.1002/ana.24999

21. Ferman TJ, Boeve BF, Smith GE, Lin SC, Silber MH et al (2011) Inclusion of RBD improves the diagnostic classification of dementia with Lewy bodies. Neurology 77:875–882. https://doi.org/10.1212/WNL.0b013e31822c9148

22. Fyfe I (2020) APOE*ε4 promotes synucleinopathy. Nat Rev Neurol 16. https://doi.org/10.1038/s41582-020-0335-5

23. Goldberg TE, Huey ED, Devanand DP (2020) Association of APOE e2 genotype with Alzheimer’s and non-Alzheimer’s neurodegenerative pathologies. Nat Commun 11. https://doi.org/10.1038/s41467-020-18198-x

24. Guerreiro R, Ross OA, Kun-Rodrigues C, Hernandez DG, Orme T et al (2018) Investigating the genetic architecture of dementia with Lewy bodies: a two-stage genome-wide association study. Lancet Neurol 17:64–74. https://doi.org/10.1016/s1474-4422(17)30400-3

25. Hohl U, Tiraboschi P, Hansen LA, Thal LJ, Corey-Bloom J (2000) Diagnostic Accuracy of Dementia With Lewy Bodies. Arch Neurol 57:347–351. https://doi.org/10.1001/archneur.57.3.347

26. Hunter JD (2007) Matplotlib: A 2D graphics environment. J Comput Inf Sci Eng 9:90–95. https://doi.org/10.1109/MCSE.2007.55

27. Kaivola K et al (2023) Genome-wide structural variant analysis identifies risk loci for non-Alzheimer’s dementias. Cell Genom 3. https://doi.org/10.1016/j.xgen.2023.100316

28. Kaivola K, Shah Z, Chia R, International LBD Genomics Consortium, Scholz S (2022) Genetic evaluation of dementia with Lewy bodies implicates distinct disease subgroups. Brain 145:1757–1762. https://doi.org/10.1093/brain/awab402

29. Karczewski KJ, Francioli LC, Tiao G et al (2020) The mutational constraint spectrum quantified from variation in 141,456 humans. Nature 581:434–443. https://doi.org/10.1038/s41586-020-2308-7

30. Kotzbauer PT, Trojanowski JQ, Lee VMY (2001) Lewy body pathology in Alzheimer’s disease. J Mol Neurosci 17:225–232. https://doi.org/10.1385/JMN:17:2:225

31. Koutsodendris N, Nelson MR, Rao A, Huang Y (2022) Apolipoprotein E and Alzheimer’s Disease: Findings, Hypotheses, and Potential Mechanisms. Annu Rev Path 17:73–99. https://doi.org/10.1146/annurev-pathmechdis-030421-112756

32. Lantero Rodriguez J, Karikari TK, Suárez-Calvet M et al (2020) Plasma p-tau181 accurately predicts Alzheimer’s disease pathology at least 8 years prior to post-mortem and improves the clinical characterisation of cognitive decline. Acta Neuropathol 140:267–278. https://doi.org/10.1007/s00401-020-02195-x

33. Le Guen Y, Belloy MЕ, Napolioni V, et al (2021) A novel age-informed approach for genetic association analysis in Alzheimer’s disease. Alzheimer’s Res Therapy 13. https://doi.org/10.1186/s13195-021-00808-5

34. Leverenz JB, Fishel MA, Peskind ER et al (2006) Lewy Body Pathology in Familial Alzheimer Disease: Evidence for Disease-and Mutation-Specific Pathologic Phenotype. Arch Neurol 63:370–376. https://doi.org/10.1001/archneur.63.3.370

35. Machiela MJ, Chanock SJ (2015) LDlink: a web-based application for exploring population-specific haplotype structure and linking correlated alleles of possible functional variants. Bioinformatics 31:3555–3557. https://doi.org/10.1093/bioinformatics/btv402

36. Majbour N, Aasly J, Abdi I, Ghanem S, Erskine D, van de Berg W, El-Agnaf O (2022) Disease-associated α-synuclein aggregates as biomarkers of Parkinson disease clinical stage. Neurology 99. https://doi.org/10.1212/WNL.0000000000201199

37. Manichaikul A, Mychaleckyj JC, Rich SS, Daly K, Sale M, Chen WM (2010) Robust relationship inference in genome-wide association studies. Bioinformatics 26:2867–2873. https://doi.org/10.1093/bioinformatics/btq559

38. McKeith IG, Boeve BF, Dickson DW, Halliday G, Taylor J et al (2017) Diagnosis and management of dementia with Lewy bodies: Fourth consensus report of the DLB Consortium. Neurology 89:88–100. https://doi.org/10.1212/WNL.0000000000004058

39. Mirra SS, Heyman A, McKeel D, Sumi SM, Crain BJ et al (1991) The Consortium to Establish a Registry for Alzheimer’s Disease (CERAD). Part II. Standardization of the neuropathologic assessment of Alzheimer’s disease. Neurology 41:479–486. https://doi.org/10.1212/wnl.41.4.479

40. Nalls MA, Blauwendraat C, Vallerga CL, Heilbron K, Bandres-Ciga S et al (2019) Identification of novel risk loci, causal insights, and heritable risk for Parkinson’s disease: a meta-analysis of genome-wide association studies. Lancet Neurol 18:1091–1102. https://doi.org/10.1016%2FS1474-4422(19)30320-5

41. Nelson PT, Alazufoff I, Bigio EH, Bouras C, Braak H et al (2012) Correlation of Alzheimer Disease Neuropathologic Changes With Cognitive Status: A Review of the Literature. J Neuropathol Exp Neurol 71:362–381. https://doi.org/10.1097/nen.0b013e31825018f7

42. Pillai JA, Bena J, Bonner-Jackson A, Leverenz JB (2021) Impact of APOE ε4 genotype on initial cognitive symptoms differs for Alzheimer’s and Lewy body neuropathology. Alzheimer’s Res Therapy 13. https://doi.org/10.1186/s13195-021-00771-1

43. Poggiolini I, Gupta V, Lawton M, Lee S, El-Turabi A et al (2022) Diagnostic value of cerebrospinal fluid alpha-synuclein seed quantification in synucleinopathies. Brain 145:584–595. https://doi.org/10.1093/brain/awab431

44. Rahimi J, Kovacs GG (2014) Prevalence of mixed pathologies in the aging brain. Alzheimer’s Res Therapy 6. https://doi.org/10.1186/s13195-014-0082-1

45. Rizzo G, Arcuti S, Copetti M, Alessandria M, Savica R et al (2018) Accuracy of clinical diagnosis of dementia with Lewy bodies: a systematic review and meta-analysis. J Neurol Neurosurg Psychiatry 89:358–366. https://doi.org/10.1136/jnnp-2017-316844

46. Robinson JL, Lee EB, Xie SX, Rennert L, Suh E et al (2018) Neurodegenerative disease concomitant proteinopathies are prevalent, age-related and APOE4-associated. Brain 141:2181–2193. https://doi.org/10.1093/brain/awy146

47. Russo MJ, Orru CD, Concha-Marambio L et al (2021) High diagnostic performance of independent alpha-synuclein seed amplification assays for detection of early Parkinson’s disease. acta neuropathol commun 9. https://doi.org/10.1186/s40478-021-01282-8

48. Sabir MS, Blauwendraat C, Ahmed S, Serrano GE, Beach TG et al (2019) Assessment of APOE in atypical parkinsonism syndromes. Neurobiol Dis 127:142–146. https://doi.org/10.1016%2Fj.nbd.2019.02.016

49. Savica R, Beach TG, Hentz JG et al (2019) Lewy body pathology in Alzheimer’s disease: A clinicopathological prospective study. Acta Neurol Scand 139:76–81. https://doi.org/10.1111/ane.13028

50. Skogseth R, Hortobágyi T, Soennesyn H, Chwiszczuk L, Ffytche D et al (2017) Accuracy of clinical diagnosis of dementia with Lewy bodies versus neuropathology. J Alzheimer’s Dis 59:1139–1152. https://doi.org/10.3233/jad-170274

51. Thal DR, Rub U, Orantes M, Braak H (2002) Phases of Aβ-deposition in the human brain and its relevance for the development of AD. Neurology 58:1791–1800. https://doi.org/10.1212/WNL.58.12.1791

52. Tsuang D, Leverenz JB, Lopez OL et al (2013) APOE ɛ4 Increases Risk for Dementia in Pure Synucleinopathies. JAMA Neurol 70:223–228. https://doi.org/10.1001/jamaneurol.2013.600

53. Twohig D, Nielsen HM (2019) α-synuclein in the pathophysiology of Alzheimer’s disease. Mol Neurodegeneration 14. https://doi.org/10.1186/s13024-019-0320-x

54. Walker JM, Richardson TE (2023) Cognitive resistance to and resilience against multiple comorbid neurodegenerative pathologies and the impact of APOE status. J Neuropathol Exp Pathol 82:110–119. https://doi.org/10.1093/jnen/nlac115

55. Willer CJ, Li Y, Abecasis GR (2010) METAL: fast and efficient meta-analysis of genomewide association scans. Bioinformatics 26:2190–2191. https://doi.org/10.1093/bioinformatics/btq340

56. Yin L, Zhang H, Tang Z, Xu J, Yin D et al (2021) rMVP: A Memory-efficient, Visualization-enhanced, and Parallel-accelerated Tool for Genome-wide Association Study. Genomics Proteomics Bioinformatics 19:619–628. https://doi.org/10.1016/j.gpb.2020.10.007

57. Zhao N et al (2020) APOE4 exacerbates alpha-synuclein pathology and related toxicity independent of amyloid. Sci Transl Med 12. https://doi.org/10.1126/scitranslmed.aay1809

