## Supplementary Materials for "*APOE*-*ε*4 and *BIN1* increase risk of Alzheimer’s disease pathology but not specifically of Lewy body pathology"

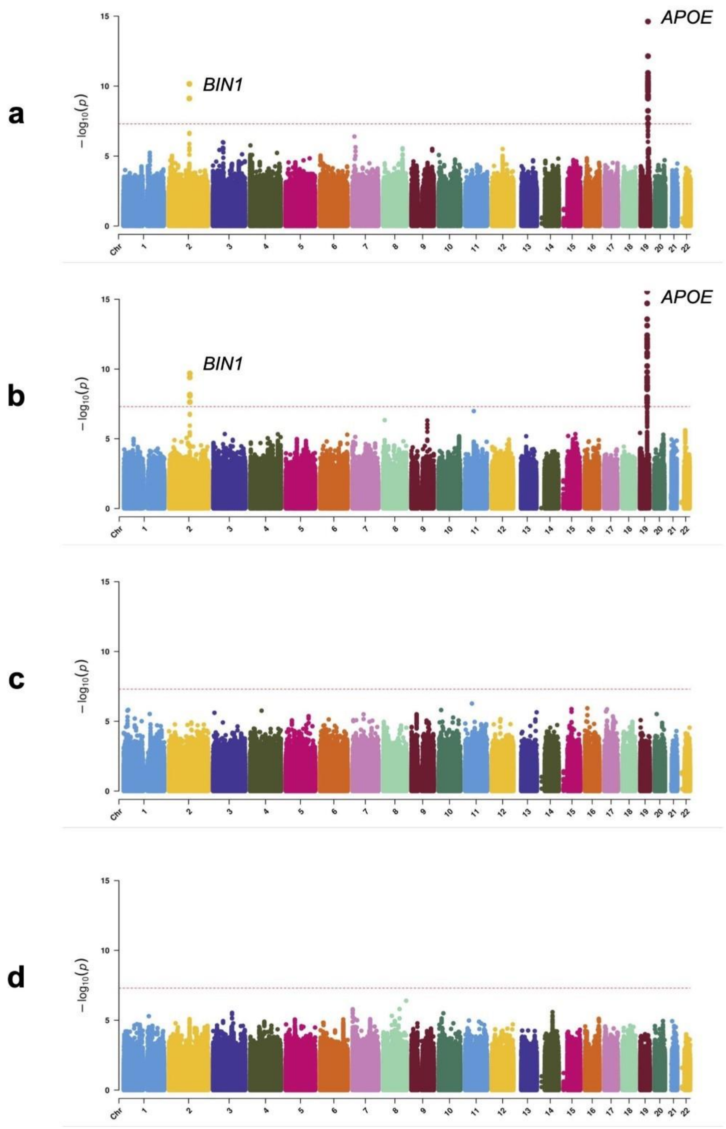


**Supplementary Figure 1. Manhattan plots of genetic association with pathology classes in the subset of NACC individuals. A.** Association with AD$^+$LB$^+$ pathology versus AD$^-$LB$^-$ pathology. **B.** Association with AD$^+$LB$^-$ pathology versus AD$^-$LB$^-$ pathology. **C.** Association with AD$^-$LB$^+$ pathology versus AD$^-$LB$^-$ pathology. **D.** Association with AD$^+$LB$^+$ pathology versus AD$^+$LB$^-$ pathology.

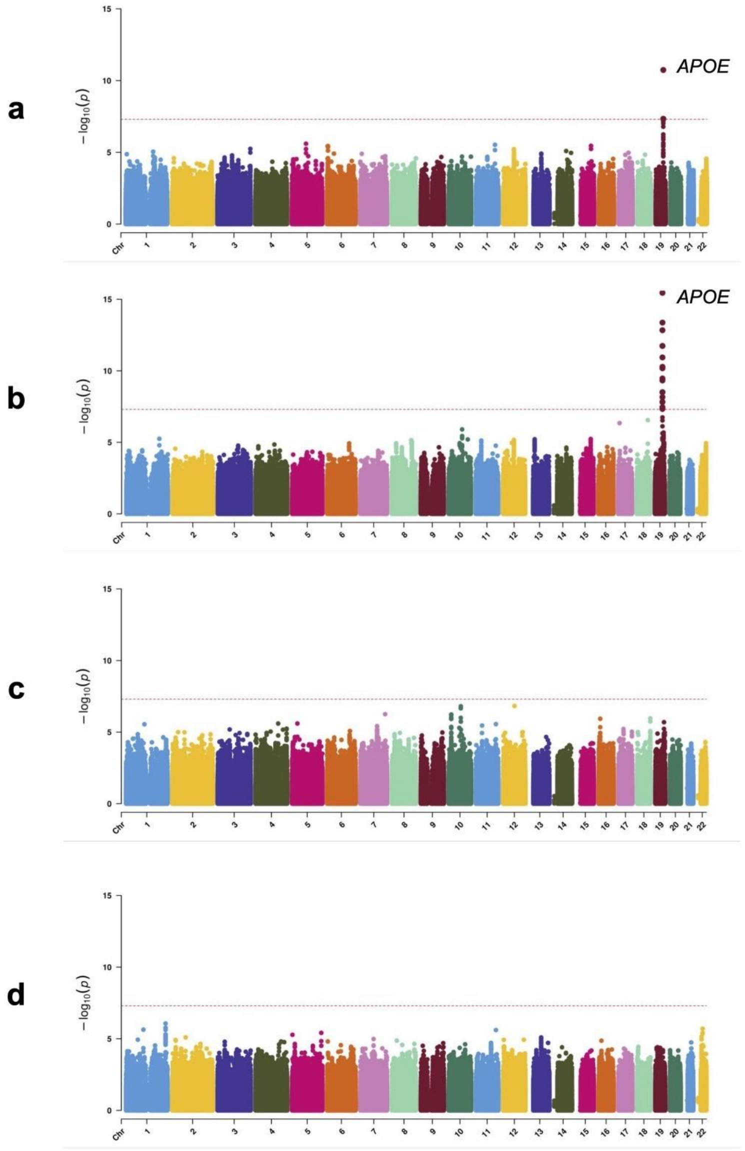


**Supplementary Figure 2. Manhattan plots of genetic association with pathology classes in the subset of Rush individuals. A.** Association with AD$^{+}$LB$^{+}$ pathology versus AD$^{-}$LB$^{-}$ pathology. **B.** Association with AD$^{+}$LB$^{-}$ pathology versus AD$^{-}$LB$^{-}$ pathology. **C.** Association with AD$^{-}$LB$^{+}$ pathology versus AD$^{-}$LB$^{-}$ pathology. **D.** Association with AD$^{+}$LB$^{+}$ pathology versus AD$^{+}$LB$^{-}$ pathology.

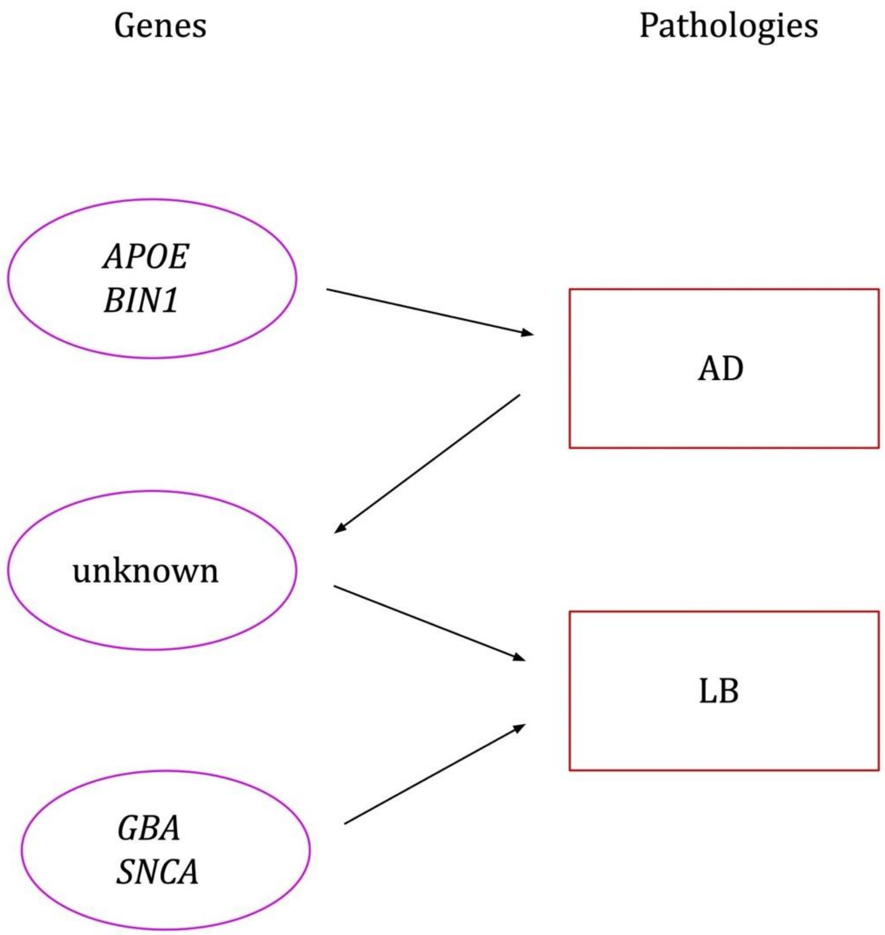


**Supplementary Figure 3. A candidate genetic model of AD and LB pathology.** *APOE* and *BIN1,* among other genes (see *e.g.,* Bellenguez *et al.*), drive AD pathology. AD pathology primes other, unknown genes to drive LB pathology. In a separate mechanism not involving AD pathology, *GBA* and *SCNA,* among other genes (see *e.g.,* Chang *et al.* (2017) and Nalls *et al.*), drive LB pathology. Notably, the set of genes including *APOE* and *BIN1* drives LB pathology only by way of AD pathology and does not affect the likelihood of LB pathology after the step of AD pathogenesis; this risk is determined by as yet unidentified genes.

**Supplementary Table 1. Comparison of pathologically assessed proportion of cohort and sample size per pathology group between reference studies [52, 2, 10, 14, 24, 18, 46, 48, 13, 28] and ours.** For Beecham *et al.,* Robinson *et al.,* and Sabir *et al.,* we consider only analyses of *APOE*-$\varepsilon$4-associated risk for LB pathology or dementia [2, 46, 48]. For Guerreiro *et al.,* we describe the larger discovery cohort [24]. If a study analyzed a cohort both with and without including certain clinically assessed subjects, we consider the analysis with the fewest clinically assessed individuals for which the effect size of *APOE*-$\varepsilon$4 and/or the top variant on *BIN1* is reported.

| Study | Total size | # subjects clinically evaluated only | # pathologically confirmed AD⁻LB⁻ | # pathologically confirmed AD⁺LB⁺ | # pathologically confirmed AD⁺LB⁻ | # pathologically confirmed AD⁻LB⁺ | # pathologically confirmed LB⁻ [a] | # pathologically confirmed LB⁺ [a] |
|---|---|---|---|---|---|---|---|---|
| **Tsuang *et al.* (2013)** | 640 | 0 (0%) | 269 | 224 | 244 | 91 | N/A | N/A |
| **Beecham *et al.* (2014)** | 3,526 | 0 (0%) | ? | ? | ? | ? | 2,391 | 1,135 |
| **Bras *et al.* (2014)** | 3,291 | 2,624 (80%) | 0 | ? | 0 | ? | 0 | 667 |
| **Chung *et al.* (2015)** | 531 | 0 (0%) | 0 | 215 | 316 | 0 | N/A | N/A |
| **Dickson *et al.* (2018)** | 1,310 | 660 (50%) | 0 | 491[b] (10+27+115+19 +111+209) | 0 | 159[b] (46+80+33) | N/A | N/A |
| **Guerreiro *et al.* (2018)** | 5,007 | 4,033 (81%) | 0 | ? | 0 | ? | 0 | 974 |
| **Robinson *et al.* (2018)** | 346 | 0 (0%) | 0 | 226[b] (19+111+10+21 +65) | 76[b] (16+60) | 44[b] (10+12+22) | N/A | N/A |
| **Sabir *et al.* (2019)** | 1,116 | 591 (53%) | 0 | ? | 0 | ? | N/A | 525 |
| **Chia *et al.* (2021)** | 6,982 | 4,588 (66%) | 605 | ? | 0 | ? | N/A | 1,789 |
| **Kaivola *et al.* (2022)** | 3,423 | 2,323–2,928[c] (68–86%) | ≤605 | 407[b] (66+341) | 0 | 88 | N/A | N/A |
| **Current sample** | 4,985 | 0 (0%) | 1,263 | 1,072 | 2,492 | 158 | N/A | N/A |

[a]These columns are populated only if an analysis was performed on a pathologically confirmed $LB^-$ or $LB^+$ group. In this case the sizes of the corresponding subgroups are marked as unknown (*e.g.,* $AD^+LB^+$ and $AD^-LB^+$ if $LB^+$ is known). [b]The given category was subdivided into multiple phenotypes, described in **Table 3,** and a separate analysis was performed on each subgroup [28, 18, 46]. [c]The control cohort of 2,928 individuals studied in Kaivola *et al.* was part of a larger set of controls studied in Chia *et al.*, of whom 605 were pathologically confirmed [28, 13]. Therefore, at most 605 of the 2,928 controls in Kaivola *et al.* were pathologically confirmed, and, accordingly, at fewest 2,323 controls were not pathologically confirmed (only clinically evaluated).

**Supplementary Table 2. Genome-wide significant loci in our study.** Chr:Pos:Minor:Major is chromosome, position (genome build hg38, GRCh38), minor allele, and major allele. MAF is minor allele frequency. [OR] [CI] (*P*) is effect size reported as odds ratio with 95% confidence interval and significance. The loci besides *BIN1* and *APOE* are novel.

| Variant | Chr:Pos:Minor:Major | Known locus or nearest gene | MAF | GWAS | OR [CI] (*P*) |
|---|---|---|---|---|---|
| **rs4663105** | 2:127133851:C:A | *BIN1* | 41.9% | $AD^+LB^+$ vs. $AD^-LB^-$ | 1.53 [1.35 1.75] (1.35e-10) |
| **rs4663105** | 2:127133851:C:A | *BIN1* | 41.9% | $AD^+LB^-$ vs. $AD^-LB^-$ | 1.40 [1.26 1.56] (6.51e-10) |
| **rs112017605** | 10:62580294:A:G | *AC024598.1/ AC067752.1* | 6.0% | $AD^-LB^+$ vs. $AD^-LB^-$ | 3.01 [2.03 4.06] (4.60e-10) |
| **rs116691607** | 17:30277568:T:C | *BLMH* | 2.4% | $AD^-LB^+$ vs. $AD^-LB^-$ | 5.17 [2.88 9.29] (3.96e-08) |
| **rs429358** | 19:44908684:C:T | *APOE* | 27.1% | $AD^+LB^+$ vs. $AD^-LB^-$ | 4.24 [3.52 5.10] (1.49e-52) |
| **rs429358** | 19:44908684:C:T | *APOE* | 27.1% | $AD^+LB^-$ vs. $AD^-LB^-$ | 4.22 [3.60 4.96] (1.41e-69) |

**Supplementary Table 3. Association of *APOE-ε2* (rs7412) with different pathology contrasts.** The first column corresponds to the current study, while the following two columns correspond to results obtained using the current sample using literature criteria to classify participants into pathology groups (**Fig. 1a–b**) [52, 28]. The last column corresponds to a result reported in the literature [48]. Effect sizes are reported as OR with 95% confidence interval [CI] and significance (*P*-value).

| | **Current sample** | **Initial sample × Tsuang *et al.* (2013) criteria** | **Initial sample × Kaivola *et al.* (2022) criteria** | **Reported by Sabir *et al.* (2019)** |
|---|---|---|---|---|
| **$AD^+LB^+$ vs. $AD^-LB^-$** | 0.39 [0.29 0.51] (3.89e-11) | 0.39 [0.29 0.54] (3.90e-09) | 0.38 [0.27 0.53] (9.24e-09) | N/A |
| **$AD^+LB^-$ vs. $AD^-LB^-$** | 0.42 [0.34 0.51] (2.68e-16) | 0.45 [0.37 0.56] (4.52e-14) | 0.44 [0.36 0.53] (9.20e-16) | N/A |
| **$AD^-LB^+$ vs. $AD^-LB^-$** | 1.17 [0.79 1.74] (4.30e-01) | 0.90 [0.65 1.24] (5.18e-01) | 1.12 [0.75 1.68] (5.72e-01) | N/A |
| **$AD^+LB^+$ vs. $AD^+LB^-$** | 0.90 [0.69 1.18] (4.55e-01) | 0.84 [0.62 1.13] (2.49e-01) | 0.85 [0.63 1.14] (2.78e-01) | N/A |
| **$LB^+$ vs. $AD^-LB^-$** | N/A | N/A | N/A | 0.39 [0.26 0.59] (6.88e-06) |

**Supplementary Table 4. Association of rs4663105 on the *BIN1* locus and *APOE-ε*4 (rs429358) with different pathology contrasts**. First two columns correspond to the current study (based on a meta-analysis) and are compared to the association in the subsets of NACC and Rush individuals separately.

| | **Current sample** | | **Current sample NACC individuals** | | **Current sample Rush individuals** | |
|---|---|---|---|---|---|---|
| | **rs4663105** | **rs429358** | **rs4663105** | **rs429358** | **rs4663105** | **rs429358** |
| **$AD^+LB^+$ vs. $AD^-LB^-$** | 1.53 [1.35 1.75] (1.35e-10) | 4.24 [3.52 5.10] (1.49e-52) | 1.68 [1.44 1.96] (7.01e-11) | 4.54 [3.66 5.63] (4.95e-43) | 1.24 [0.98 1.57] (7.88e-02) | 3.48 [2.42 5.01] (1.83e-11) |
| **$AD^+LB^-$ vs. $AD^-LB^-$** | 1.40 [1.26 1.56] (6.51e-10) | 4.22 [3.60 4.96] (1.41e-69) | 1.52 [1.33 1.73] (3.96e-10) | 4.54 [3.66 5.63] (4.95e-43) | 1.19 [0.99 1.43] (6.43e-02) | 3.53 [2.60 4.77] (3.56e-16) |
| **$AD^-LB^+$ vs. $AD^-LB^-$** | 1.10 [0.85 1.41] (4.76e-01) | 0.93 [0.60 1.43] (7.34e-01) | 1.01 [0.72 1.42] (9.57e-01) | 1.05 [0.62 1.78] (8.53e-01) | 1.20 [0.83 1.72] (3.27e-01) | 0.72 [0.34 1.53] (3.94e-01) |
| **$AD^+LB^+$ vs. $AD^+LB^-$** | 1.13 [1.02 1.25] (1.91e-02) | 1.01 [0.90 1.13] (8.33e-01) | 1.15 [1.03 1.30] (1.38e-02) | 1.01 [0.90 1.14] (8.36e-01) | 1.04 [0.82 1.31] (7.72e-01) | 1.01 [0.74 1.38] (9.60e-01) |

**Supplementary Table 5. Associations of known Alzheimer's disease risk loci reported in Bellenguez *et al.* (2022) and known Parkinson's disease risk loci reported in Chang *et al.* (2017) and Nalls *et al.* (2019) with different pathology contrasts** [3, 12, 40]. Loci for which the reported variant was missing from our summary statistics are not included. Chr:Pos:Minor:Major is chromosome, position, and minor allele and major allele in our study. Lit. MAF is the frequency reported in the literature of the minor allele in our study. Lit. effect is the effect size reported in the literature. N+R MAF is the minor allele frequency in our study. Nominally significant associations ($P < 0.05$) are colored red (minor allele increases risk) and blue (minor allele decreases risk), associations with $P < 0.10$ are colored green regardless of the direction of effect.

| Variant | Chr:Pos:Minor:Major | Locus | Lit. MAF | Lit. effect | Study | N+R MAF | AD$^+$LB$^+$ vs. AD$^-$LB$^-$ | AD$^+$LB$^-$ vs. AD$^-$LB$^-$ | AD$^-$LB$^+$ vs. AD$^-$LB$^-$ | AD$^+$LB$^+$ vs. AD$^+$LB$^-$ |
|---|---|---|---|---|---|---|---|---|---|---|
| **rs35749011** | 1:155162560:A:G | *GBA* | 2.4% | 1.72 [1.59 1.89] (2.59e-35) | Chang *et al.* (2017) | 1.3% | 1.60 [0.87 2.91] (1.28e-01) | 0.82 [0.47 1.44] (4.89e-01) | 2.10 [0.74 5.93] (1.62e-01) | 1.94 [1.24 3.06] (4.04e-03) |
| **rs6658353** | 1:161499264:C:G | *FCGR2A* | 50.1% | 1.07 [1.05 1.09] (6.10e-12) | Nalls *et al.* (2019) | 49.5% | 0.94 [0.82 1.07] (3.53e-01) | 0.97 [0.87 1.08] (5.51e-01) | 0.89 [0.68 1.15] (3.64e-01) | 0.99 [0.89 1.10] (8.32e-01) |
| **rs11578699** | 1:171750629:T:C | *VAMP4* | 19.5% | 0.93 [0.91 0.95] (4.47e-09) | Nalls *et al.* (2019) | 19.3% | 1.04 [0.88 1.22] (6.48e-01) | 0.98 [0.86 1.11] (7.18e-01) | 1.20 [0.90 1.60] (2.22e-01) | 1.06 [0.93 1.20] (4.05e-01) |
| **rs823118** | 1:205754444:C:T | *NUCKS1/ SLC41A1* | 46.7% | 0.89 [0.87 0.91] (1.12e-23) | Chang *et al.* (2017) | 44.4% | 0.94 [0.83 1.07] (3.34e-01) | 0.94 [0.85 1.05] (2.63e-01) | 1.05 [0.82 1.34] (7.03e-01) | 0.97 [0.88 1.08] (6.04e-01) |
| **rs679515** | 1:207577223:T:C | *CR1* | 18.8% | 1.13 [1.11 1.15] (7.20e-46) | Bellenguez *et al.* (2022) | 20.2% | 1.09 [0.93 1.29] (2.98e-01) | 1.19 [1.04 1.36] (9.99e-03) | 0.91 [0.66 1.25] (5.55e-01) | 0.95 [0.83 1.08] (4.22e-01) |
| **rs4653767** | 1:226728377:C:T | *ITPKB* | 31.5% | 0.92 [0.90 0.94] (1.63e-11) | Chang *et al.* (2017) | 29.2% | 0.92 [0.80 1.06] (2.43e-01) | 0.94 [0.84 1.06] (3.23e-01) | 0.85 [0.65 1.12] (2.47e-01) | 0.95 [0.85 1.06] (3.38e-01) |
| **rs10797576** | 1:232528865:T:C | *SIPA1L2* | 13.7% | 1.12 [1.09 1.15] (8.41e-13) | Chang *et al.* (2017) | 13.2% | 1.04 [0.87 1.26] (6.39e-01) | 1.05 [0.90 1.22] (5.34e-01) | 1.18 [0.84 1.65] (3.46e-01) | 1.00 [0.86 1.17] (9.63e-01) |
| **rs72777026** | 2:9558882:G:A | *ADAM17* | 14.4% | 1.06 [1.04 1.08] (2.70e-08) | Bellenguez *et al.* (2022) | 14.2% | 1.30 [1.09 1.56] (3.92e-03) | 1.17 [1.01 1.37] (4.19e-02) | 1.44 [1.04 2.00] (2.81e-02) | 1.13 [0.98 1.31] (8.13e-02) |
| **rs76116224** | 2:17966582:T:A | *KCNS3* | 9.6% | 1.12 [1.08 1.16] (1.27e-08) | Nalls *et al.* (2019) | 9.7% | 0.95 [0.77 1.18] (6.57e-01) | 0.94 [0.79 1.12] (4.83e-01) | 1.05 [0.71 1.55] (8.03e-01) | 1.06 [0.89 1.27] (5.14e-01) |
| **rs17020490** | 2:37304796:C:T | *PRKD3* | 14.5% | 1.06 [1.04 1.08] (3.30e-09) | Bellenguez *et al.* (2022) | 14.6% | 1.10 [0.91 1.31] (3.26e-01) | 1.04 [0.90 1.21] (5.81e-01) | 1.07 [0.74 1.54] (7.25e-01) | 1.04 [0.90 1.19] (6.19e-01) |
| **rs2042477** | 2:95335195:A:T | *KCNIP3* | 24.2% | 0.94 [0.92 0.96] (1.38e-08) | Nalls *et al.* (2019) | 24.7% | 0.95 [0.82 1.11] (5.33e-01) | 0.90 [0.79 1.01] (8.34e-02) | 1.00 [0.76 1.32] (9.88e-01) | 1.06 [0.94 1.20] (3.56e-01) |
| **rs34043159** | 2:101796654:C:T | *IL1R2* | 35.2% | 1.08 [1.06 1.10] (5.48e-11) | Chang *et al.* (2017) | 33.1% | 1.00 [0.87 1.15] (9.66e-01) | 0.96 [0.86 1.08] (5.06e-01) | 1.08 [0.83 1.41] (5.59e-01) | 1.04 [0.93 1.16] (5.30e-01) |
| **rs6733839** | 2:127135234:T:C | *BIN1* | 38.9% | 1.17 [1.16 1.19] (6.10e-118) | Bellenguez *et al.* (2022) | 41.9% | 1.52 [1.34 1.74] (4.60e-10) | 1.43 [1.29 1.60] (7.49e-11) | 1.12 [0.87 1.45] (3.72e-01) | 1.10 [0.99 1.22] (6.57e-02) |
| **rs6430538** | 2:134782397:T:C | *TMEM163/ CCNT2* | 48.8% | 0.89 [0.87 0.91] (8.24e-24) | Chang *et al.* (2017) | 43.7% | 1.01 [0.89 1.15] (8.36e-01) | 1.12 [1.00 1.24] (4.01e-02) | 0.86 [0.67 1.10] (2.27e-01) | 0.91 [0.82 1.01] (8.31e-02) |

| | | | | | | | | | | |
|---|---|---|---|---|---|---|---|---|---|---|
| **rs353116** | 2:165277122:T:C | *SCN3A* | 38.5% | 0.94 [0.92 0.96] (2.98e-08) | Chang *et al.* (2017) | 38.7% | 0.85 [0.74 0.96] (1.14e-02) | 0.94 [0.84 1.04] (2.46e-01) | 0.90 [0.70 1.15] (4.02e-01) | 0.95 [0.86 1.06] (3.52e-01) |
| **rs1474055** | 2:168253884:T:C | *STK39* | 11.9% | 1.20 [1.16 1.25] (5.68e-26) | Chang *et al.* (2017) | 12.2% | 0.85 [0.70 1.04] (1.17e-01) | 1.00 [0.86 1.17] (9.74e-01) | 0.96 [0.66 1.39] (8.16e-01) | 0.88 [0.75 1.03] (1.11e-01) |
| **rs139643391** | 2:202878716:T:TC | *WDR12* | 13.1% | 0.94 [0.92 0.96] (1.10e-08) | Bellenguez *et al.* (2022) | 12.2% | 0.97 [0.80 1.16] (7.11e-01) | 0.94 [0.80 1.10] (4.33e-01) | 1.15 [0.83 1.61] (4.04e-01) | 0.98 [0.84 1.15] (8.22e-01) |
| **rs10933431** | 2:233117202:G:C | *INPP5D* | 23.4% | 0.93 [0.92 0.95] (3.60e-18) | Bellenguez *et al.* (2022) | 21.8% | 0.87 [0.74 1.01] (7.37e-02) | 0.79 [0.70 0.90] (3.09e-04) | 1.09 [0.83 1.44] (5.42e-01) | 1.07 [0.94 1.22] (2.99e-01) |
| **rs4073221** | 3:18235996:G:T | *SATB1* | 13.2% | 1.10 [1.06 1.13] (1.57e-08) | Chang *et al.* (2017) | 12.9% | 1.04 [0.86 1.26] (6.68e-01) | 1.08 [0.92 1.26] (3.55e-01) | 0.97 [0.67 1.39] (8.65e-01) | 1.03 [0.88 1.19] (7.40e-01) |
| **rs6808178** | 3:28664199:T:C | *LINC00693* | 37.9% | 1.07 [1.05 1.09] (8.09e-12) | Nalls *et al.* (2019) | 37.5% | 0.90 [0.79 1.03] (1.27e-01) | 0.99 [0.89 1.10] (8.79e-01) | 0.84 [0.65 1.07] (1.64e-01) | 0.90 [0.81 1.01] (6.81e-02) |
| **rs12497850** | 3:48711556:G:T | *NCKIPSD/ CDC71* | 34.7% | 0.93 [0.91 0.96] (9.16e-09) | Chang *et al.* (2017) | 34.8% | 1.00 [0.87 1.14] (9.60e-01) | 1.04 [0.93 1.16] (4.95e-01) | 1.09 [0.84 1.41] (5.30e-01) | 0.98 [0.87 1.09] (6.53e-01) |
| **rs115185635** | 3:87471707:C:G | *CHMP2B* | 3.6% | 1.21 [1.10 1.33] (1.22e-04) | Chang *et al.* (2017) | 3.9% | 0.97 [0.69 1.34] (8.33e-01) | 1.03 [0.79 1.34] (8.36e-01) | 0.79 [0.40 1.56] (5.06e-01) | 0.92 [0.70 1.21] (5.62e-01) |
| **rs55961674** | 3:122478045:T:C | *KPNA1* | 17.2% | 1.09 [1.06 1.12] (9.98e-12) | Nalls *et al.* (2019) | 16.9% | 0.97 [0.82 1.15] (7.47e-01) | 1.04 [0.91 1.20] (5.47e-01) | 0.95 [0.68 1.32] (7.59e-01) | 0.97 [0.85 1.12] (6.92e-01) |
| **rs11707416** | 3:151391177:A:T | *MED12L* | 36.7% | 0.94 [0.92 0.96] (1.13e-10) | Nalls *et al.* (2019) | 37.3% | 1.00 [0.88 1.14] (9.93e-01) | 1.07 [0.96 1.19] (2.47e-01) | 1.04 [0.81 1.32] (7.83e-01) | 0.95 [0.86 1.06] (3.94e-01) |
| **rs16824536** | 3:155069722:A:G | *MME* | 5.4% | 0.92 [0.89 0.95] (3.60e-08) | Bellenguez *et al.* (2022) | 5.4% | 0.80 [0.61 1.04] (9.94e-02) | 0.77 [0.62 0.96] (2.09e-02) | 1.22 [0.78 1.93] (3.86e-01) | 1.10 [0.87 1.40] (4.12e-01) |
| **rs61762319** | 3:155084189:G:A | *MME* | 2.6% | 1.16 [1.11 1.21] (2.20e-11) | Bellenguez *et al.* (2022) | 2.6% | 0.97 [0.64 1.48] (8.97e-01) | 1.32 [0.95 1.82] (9.44e-02) | 0.66 [0.26 1.70] (3.87e-01) | 0.72 [0.52 1.00] (5.34e-02) |
| **rs1450522** | 3:161359842:G:A | *SPTSSB* | 32.6% | 1.06 [1.04 1.09] (5.01e-10) | Nalls *et al.* (2019) | 32.0% | 0.92 [0.81 1.06] (2.38e-01) | 0.96 [0.86 1.08] (5.25e-01) | 1.03 [0.80 1.33] (8.15e-01) | 0.96 [0.86 1.07] (4.19e-01) |
| **rs12637471** | 3:183044649:A:G | *MCCC1* | 21.9% | 0.85 [0.82 0.87] (2.11e-30) | Chang *et al.* (2017) | 20.2% | 1.03 [0.88 1.20] (7.21e-01) | 0.94 [0.83 1.07] (3.35e-01) | 0.85 [0.63 1.15] (2.93e-01) | 1.07 [0.94 1.21] (3.30e-01) |
| **rs34311866** | 4:958159:C:T | *TMEM175/ DGKQ* | 19.9% | 1.23 [1.20 1.27] (1.47e-50) | Chang *et al.* (2017) | 19.1% | 1.07 [0.91 1.24] (4.24e-01) | 0.89 [0.78 1.02] (9.56e-02) | 1.33 [1.01 1.75] (4.29e-02) | 1.22 [1.07 1.38] (2.83e-03) |
| **rs6846529** | 4:11023507:C:T | *CLNK/ HS3ST1* | 28.3% | 1.07 [1.05 1.08] (2.20e-17) | Bellenguez *et al.* (2022) | 28.0% | 1.11 [0.96 1.28] (1.46e-01) | 1.15 [1.02 1.29] (2.12e-02) | 1.01 [0.77 1.32] (9.49e-01) | 0.97 [0.86 1.08] (5.66e-01) |
| **rs11724635** | 4:15735478:C:A | *FAM200B/ CD38* | 43.7% | 0.90 [0.88 0.92] (1.22e-19) | Chang *et al.* (2017) | 45.3% | 1.01 [0.89 1.15] (8.64e-01) | 0.97 [0.87 1.08] (5.37e-01) | 0.71 [0.55 0.92] (9.03e-03) | 1.05 [0.95 1.17] (3.28e-01) |
| **rs34025766** | 4:17967188:A:T | *LCORL* | 15.9% | 0.92 [0.90 0.94] (2.87e-10) | Nalls *et al.* (2019) | 16.4% | 1.15 [0.97 1.36] (1.11e-01) | 1.04 [0.90 1.20] (5.81e-01) | 0.92 [0.65 1.29] (6.25e-01) | 1.10 [0.96 1.26] (1.84e-01) |
| **rs2245466** | 4:40197226:G:C | *RHOH* | 34.3% | 1.05 [1.03 1.06] (1.20e-09) | Bellenguez *et al.* (2022) | 33.2% | 1.06 [0.93 1.22] (3.74e-01) | 1.04 [0.93 1.17] (5.05e-01) | 0.97 [0.74 1.27] (8.27e-01) | 1.04 [0.93 1.16] (5.13e-01) |
| **rs6812193** | 4:76277833:T:C | *FAM47E* | 39.8% | 0.92 [0.90 0.94] (1.43e-14) | Chang *et al.* (2017) | 37.0% | 0.91 [0.80 1.03] (1.44e-01) | 0.98 [0.88 1.09] (7.55e-01) | 0.92 [0.72 1.18] (5.00e-01) | 0.95 [0.85 1.05] (3.06e-01) |

| | | | | | | | | | | |
|---|---|---|---|---|---|---|---|---|---|---|
| **rs356182** | 4:89704960:G:A | *SNCA* | 37.5% | 0.75 [0.74 0.77] (5.21e-123) | Chang *et al.* (2017) | 34.9% | 0.90 [0.79 1.03] (1.37e-01) | 0.95 [0.85 1.07] (4.04e-01) | 0.74 [0.57 0.97] (2.60e-02) | 0.97 [0.87 1.08] (5.38e-01) |
| **rs78738012** | 4:113439216:C:T | *ANK2/ CAMK2D* | 10.6% | 1.13 [1.09 1.17] (4.78e-11) | Chang *et al.* (2017) | 10.0% | 1.05 [0.86 1.30] (6.18e-01) | 1.00 [0.84 1.19] (9.78e-01) | 1.07 [0.72 1.61] (7.27e-01) | 1.02 [0.86 1.21] (8.27e-01) |
| **rs62333164** | 4:169662006:A:G | *CLCN3* | 32.6% | 0.94 [0.92 0.96] (2.00e-10) | Nalls *et al.* (2019) | 32.7% | 1.06 [0.93 1.21] (3.73e-01) | 1.14 [1.02 1.28] (1.74e-02) | 1.14 [0.88 1.46] (3.25e-01) | 0.96 [0.86 1.07] (4.79e-01) |
| **rs112403360** | 5:14724304:A:T | *ANKH* | 7.3% | 1.09 [1.06 1.12] (2.30e-09) | Bellenguez *et al.* (2022) | 7.4% | 1.05 [0.81 1.36] (7.20e-01) | 1.30 [1.06 1.59] (1.13e-02) | 1.24 [0.80 1.93] (3.37e-01) | 0.82 [0.67 1.00] (5.05e-02) |
| **rs2694528** | 5:60978096:C:A | *ELOVL7* | 11.5% | 1.15 [1.11 1.20] (4.84e-15) | Chang *et al.* (2017) | 8.5% | 0.89 [0.71 1.12] (3.13e-01) | 0.93 [0.78 1.12] (4.41e-01) | 0.76 [0.47 1.23] (2.69e-01) | 1.01 [0.83 1.22] (9.46e-01) |
| **rs62374257** | 5:86927378:C:T | *COX7C* | 23.0% | 1.07 [1.05 1.09] (1.40e-15) | Bellenguez *et al.* (2022) | 22.8% | 1.21 [1.04 1.41] (1.63e-02) | 1.26 [1.11 1.43] (3.83e-04) | 1.17 [0.87 1.57] (3.09e-01) | 0.96 [0.85 1.08] (5.03e-01) |
| **rs26431** | 5:103030090:G:C | *PAM* | 29.7% | 0.94 [0.92 0.96] (1.57e-09) | Nalls *et al.* (2019) | 30.6% | 1.01 [0.88 1.16] (9.07e-01) | 1.00 [0.89 1.12] (9.32e-01) | 1.09 [0.83 1.42] (5.41e-01) | 1.05 [0.94 1.18] (3.70e-01) |
| **rs11950533** | 5:134863415:A:C | *C5orf24* | 10.2% | 0.91 [0.88 0.94] (7.16e-09) | Nalls *et al.* (2019) | 10.4% | 0.84 [0.68 1.02] (8.48e-02) | 0.83 [0.70 0.98] (2.70e-02) | 0.72 [0.47 1.10] (1.33e-01) | 0.96 [0.81 1.14] (6.48e-01) |
| **rs871269** | 5:151052827:T:C | *TNIP1* | 32.6% | 0.96 [0.95 0.97] (8.70e-09) | Bellenguez *et al.* (2022) | 32.4% | 0.95 [0.83 1.09] (4.59e-01) | 0.95 [0.85 1.06] (3.78e-01) | 0.99 [0.77 1.29] (9.67e-01) | 1.03 [0.93 1.15] (5.70e-01) |
| **rs113706587** | 5:180201150:A:G | *RASGEF1C* | 11.0% | 1.09 [1.07 1.12] (2.20e-16) | Bellenguez *et al.* (2022) | 10.8% | 1.09 [0.89 1.34] (4.03e-01) | 1.12 [0.95 1.33] (1.85e-01) | 0.92 [0.61 1.39] (6.95e-01) | 0.96 [0.81 1.13] (6.00e-01) |
| **rs9468199** | 6:27713436:A:G | *ZNF184* | 17.2% | 1.11 [1.08 1.14] (1.46e-12) | Chang *et al.* (2017) | 17.8% | 1.07 [0.91 1.27] (4.16e-01) | 1.00 [0.87 1.14] (9.80e-01) | 1.02 [0.74 1.41] (9.14e-01) | 1.06 [0.93 1.21] (3.77e-01) |
| **rs9261484** | 6:30140906:T:C | *TRIM40* | 24.5% | 0.94 [0.92 0.96] (1.62e-08) | Nalls *et al.* (2019) | 25.1% | 1.08 [0.94 1.25] (2.75e-01) | 1.06 [0.94 1.19] (3.46e-01) | 1.01 [0.77 1.33] (9.40e-01) | 1.01 [0.90 1.14] (8.47e-01) |
| **rs6605556** | 6:32615322:G:A | *HLA* | 16.1% | 0.91 [0.90 0.93] (7.10e-20) | Bellenguez *et al.* (2022) | 16.0% | 0.81 [0.69 0.96] (1.67e-02) | 0.87 [0.76 1.00] (4.57e-02) | 1.01 [0.74 1.38] (9.43e-01) | 0.97 [0.84 1.12] (6.81e-01) |
| **rs9275326** | 6:32698883:T:C | *HLA-DRB6/ HLA-DQA1* | 11.4% | 0.85 [0.82 0.89] (1.26e-13) | Chang *et al.* (2017) | 9.8% | 0.74 [0.60 0.91] (4.75e-03) | 0.83 [0.70 0.98] (2.84e-02) | 0.97 [0.66 1.44] (8.86e-01) | 0.88 [0.74 1.06] (1.74e-01) |
| **rs10947943** | 6:41036354:A:G | *TREM2* | 14.2% | 0.94 [0.93 0.96] (1.10e-09) | Bellenguez *et al.* (2022) | 14.4% | 0.89 [0.75 1.07] (2.06e-01) | 0.86 [0.75 1.00] (4.51e-02) | 1.05 [0.76 1.46] (7.71e-01) | 1.06 [0.91 1.22] (4.51e-01) |
| **rs143332484** | 6:41161469:T:C | *TREM2* | 1.3% | 1.41 [1.32 1.50] (2.80e-25) | Bellenguez *et al.* (2022) | 1.3% | 1.99 [1.09 3.62] (2.50e-02) | 1.87 [1.11 3.16] (1.97e-02) | 1.69 [0.55 5.18] (3.60e-01) | 1.35 [0.89 2.04] (1.58e-01) |
| **rs7767350** | 6:47517390:T:C | *CD2AP* | 27.1% | 1.08 [1.06 1.09] (7.90e-22) | Bellenguez *et al.* (2022) | 28.2% | 1.12 [0.97 1.29] (1.18e-01) | 1.12 [0.99 1.25] (6.63e-02) | 0.98 [0.74 1.28] (8.69e-01) | 0.99 [0.88 1.11] (8.49e-01) |
| **rs12528068** | 6:71778059:T:C | *RIMS1* | 28.4% | 1.07 [1.05 1.09] (1.63e-10) | Nalls *et al.* (2019) | 27.9% | 1.02 [0.89 1.18] (7.55e-01) | 1.10 [0.98 1.24] (9.40e-02) | 0.94 [0.72 1.24] (6.83e-01) | 0.96 [0.85 1.07] (4.36e-01) |
| **rs997368** | 6:111922088:G:A | *FYN* | 19.5% | 0.93 [0.91 0.95] (1.84e-09) | Nalls *et al.* (2019) | 19.6% | 0.92 [0.78 1.08] (3.11e-01) | 0.92 [0.78 1.08] (3.11e-01) | 0.92 [0.78 1.08] (3.11e-01) | 0.88 [0.77 1.01] (6.21e-02) |
| **rs785129** | 6:114291731:T:C | *HS3ST5* | 35.0% | 1.04 [1.03 1.06] (2.40e-09) | Bellenguez *et al.* (2022) | 34.7% | 1.19 [1.04 1.36] (1.25e-02) | 1.11 [0.99 1.24] (7.66e-02) | 1.13 [0.87 1.47] (3.41e-01) | 1.05 [0.94 1.17] (4.25e-01) |
| **rs75859381** | 6:132889222:C:T | *RPS12* | 3.3% | 1.25 [1.16 1.33] (1.04e-10) | Nalls *et al.* (2019) | 3.1% | 0.83 [0.58 1.19] (3.07e-01) | 0.80 [0.60 1.08] (1.48e-01) | 1.08 [0.54 2.20] (8.21e-01) | 1.13 [0.85 1.52] (3.99e-01) |

| | | | | | | | | | | |
|---|---|---|---|---|---|---|---|---|---|---|
| **rs6943429** | 7:7817263:T:C | *UMAD1* | 42.0% | 1.05 [1.03 1.06] (1.00e-10) | Bellenguez *et al.* (2022) | 42.6% | 1.00 [0.88 1.15] (9.65e-01) | 1.08 [0.97 1.21] (1.44e-01) | 0.83 [0.65 1.07] (1.52e-01) | 0.91 [0.82 1.02] (9.94e-02) |
| **rs10952097** | 7:8204382:T:C | *ICA1* | 11.4% | 1.07 [1.05 1.10] (6.80e-09) | Bellenguez *et al.* (2022) | 10.0% | 1.08 [0.87 1.34] (4.67e-01) | 1.10 [0.93 1.31] (2.73e-01) | 0.80 [0.51 1.24] (3.09e-01) | 1.01 [0.85 1.20] (9.14e-01) |
| **rs13237518** | 7:12229967:A:C | *TMEM106B* | 41.1% | 0.96 [0.94 0.97] (4.90e-11) | Bellenguez *et al.* (2022) | 42.1% | 1.23 [1.08 1.39] (1.29e-03) | 1.12 [1.01 1.24] (3.34e-02) | 1.22 [0.97 1.55] (9.14e-02) | 1.05 [0.95 1.17] (3.09e-01) |
| **rs199347** | 7:23254127:G:A | *KLHL7/ NUPL2/ GPNMB* | 36.8% | 0.91 [0.89 0.93] (3.51e-18) | Chang *et al.* (2017) | 41.5% | 1.08 [0.95 1.23] (2.39e-01) | 1.13 [1.01 1.25] (2.57e-02) | 1.04 [0.81 1.32] (7.74e-01) | 0.92 [0.83 1.03] (1.40e-01) |
| **rs1160871** | 7:28129126:G:GTCTT | *JAZF1* | 22.2% | 0.95 [0.93 0.97] (9.80e-09) | Bellenguez *et al.* (2022) | 21.9% | 0.92 [0.79 1.07] (2.70e-01) | 0.97 [0.86 1.10] (6.46e-01) | 1.07 [0.80 1.42] (6.47e-01) | 0.98 [0.86 1.11] (7.24e-01) |
| **rs6966331** | 7:37844191:T:C | *NME8* | 34.9% | 0.96 [0.94 0.97] (4.60e-10) | Bellenguez *et al.* (2022) | 33.5% | 1.03 [0.90 1.18] (6.75e-01) | 0.91 [0.82 1.02] (1.11e-01) | 0.98 [0.76 1.25] (8.56e-01) | 1.11 [0.99 1.24] (7.40e-02) |
| **rs76928645** | 7:54873635:T:C | *SEC61G* | 10.3% | 0.93 [0.91 0.95] (1.60e-10) | Bellenguez *et al.* (2022) | 10.2% | 0.81 [0.65 1.00] (4.84e-02) | 0.90 [0.76 1.07] (2.33e-01) | 1.10 [0.76 1.59] (6.02e-01) | 0.92 [0.77 1.10] (3.59e-01) |
| **rs76949143** | 7:66544864:A:T | *GS1-124K5.11* | 5.1% | 0.87 [0.82 0.91] (1.43e-08) | Nalls *et al.* (2019) | 4.7% | 1.09 [0.81 1.45] (5.77e-01) | 0.98 [0.77 1.25] (8.99e-01) | 0.90 [0.49 1.64] (7.34e-01) | 1.08 [0.85 1.37] (5.32e-01) |
| **rs7384878** | 7:100334426:C:T | *ZCWPW1/ NYAP1* | 31.0% | 0.92 [0.91 0.94] (1.10e-26) | Bellenguez *et al.* (2022) | 29.3% | 0.95 [0.82 1.09] (4.43e-01) | 0.85 [0.75 0.95] (5.45e-03) | 1.26 [0.97 1.63] (8.23e-02) | 0.94 [0.84 1.05] (2.67e-01) |
| **rs11771145** | 7:143413669:A:G | *EPHA1* | 34.8% | 0.95 [0.93 0.96] (3.30e-14) | Bellenguez *et al.* (2022) | 34.0% | 0.95 [0.84 1.09] (4.84e-01) | 0.92 [0.82 1.02] (1.26e-01) | 1.23 [0.96 1.57] (1.01e-01) | 1.02 [0.92 1.14] (7.14e-01) |
| **rs1065712** | 8:11844613:C:G | *CTSB* | 5.3% | 1.09 [1.06 1.12] (1.90e-09) | Bellenguez *et al.* (2022) | 5.8% | 1.34 [1.03 1.75] (3.22e-02) | 1.09 [0.87 1.38] (4.46e-01) | 0.85 [0.48 1.49] (5.67e-01) | 1.23 [1.00 1.52] (5.43e-02) |
| **rs2740594** | 8:11849665:G:A | *CTSB* | 24.7% | 0.92 [0.89 0.93] (5.91e-12) | Chang *et al.* (2017) | 27.0% | 1.11 [0.96 1.28] (1.47e-01) | 1.05 [0.93 1.18] (4.01e-01) | 0.93 [0.71 1.23] (6.08e-01) | 1.01 [0.90 1.13] (8.84e-01) |
| **rs591323** | 8:16839582:A:G | *MICU3* | 29.3% | 0.91 [0.89 0.94] (2.38e-11) | Chang *et al.* (2017) | 27.2% | 1.18 [1.02 1.36] (2.28e-02) | 1.04 [0.92 1.17] (5.58e-01) | 1.11 [0.85 1.45] (4.27e-01) | 1.08 [0.97 1.22] (1.69e-01) |
| **rs2280104** | 8:22668467:T:C | *SORBS3/ PDLIM2/ C8orf58/ BIN3* | 36.7% | 1.07 [1.04 1.09] (2.53e-08) | Chang *et al.* (2017) | 35.3% | 0.97 [0.85 1.12] (6.98e-01) | 1.03 [0.92 1.15] (6.12e-01) | 1.10 [0.85 1.41] (4.78e-01) | 0.95 [0.85 1.06] (3.43e-01) |
| **rs73223431** | 8:27362470:T:C | *PTK2B* | 36.9% | 1.07 [1.06 1.08] (4.00e-22) | Bellenguez *et al.* (2022) | 36.4% | 1.13 [0.99 1.29] (7.98e-02) | 1.16 [1.04 1.29] (6.67e-03) | 1.23 [0.96 1.58] (1.04e-01) | 1.00 [0.90 1.11] (9.94e-01) |
| **rs11787077** | 8:27607795:T:C | *CLU* | 39.2% | 0.91 [0.90 0.92] (1.70e-44) | Bellenguez *et al.* (2022) | 38.5% | 0.85 [0.74 0.97] (1.33e-02) | 0.89 [0.80 0.99] (3.43e-02) | 0.98 [0.77 1.26] (8.81e-01) | 0.95 [0.85 1.06] (3.62e-01) |
| **rs2086641** | 8:129889663:C:T | *FAM49B* | 27.7% | 1.06 [1.04 1.09] (1.81e-08) | Nalls *et al.* (2019) | 27.5% | 0.97 [0.84 1.12] (6.69e-01) | 1.00 [0.89 1.13] (9.38e-01) | 1.23 [0.94 1.62] (1.32e-01) | 1.03 [0.92 1.16] (5.74e-01) |
| **rs34173062** | 8:144103704:A:G | *SHARPIN* | 8.1% | 1.13 [1.09 1.16] (1.70e-16) | Bellenguez *et al.* (2022) | 8.3% | 0.95 [0.75 1.20] (6.48e-01) | 1.12 [0.93 1.35] (2.34e-01) | 1.31 [0.89 1.95] (1.76e-01) | 0.87 [0.72 1.06] (1.66e-01) |
| **rs13294100** | 9:17579692:T:G | *SH3GL2* | 37.1% | 0.92 [0.89 0.94] (4.84e-13) | Chang *et al.* (2017) | 35.5% | 1.00 [0.87 1.14] (9.93e-01) | 0.94 [0.84 1.05] (2.74e-01) | 0.93 [0.72 1.20] (5.82e-01) | 1.04 [0.93 1.15] (5.14e-01) |
| **rs6476434** | 9:34046393:C:T | *UBAP2* | 26.6% | 1.06 [1.04 1.09] (6.58e-09) | Nalls *et al.* (2019) | 27.1% | 1.02 [0.88 1.18] (7.94e-01) | 0.99 [0.88 1.11] (8.28e-01) | 0.88 [0.66 1.16] (3.51e-01) | 1.05 [0.93 1.18] (4.15e-01) |

| | | | | | | | | | | |
|---|---|---|---|---|---|---|---|---|---|---|
| **rs1800978** | 9:104903697:G:C | *ABCA1* | 13.0% | 1.06 [1.04 1.08] (1.60e-09) | Bellenguez *et al.* (2022) | 13.8% | 1.02 [0.85 1.22] (8.18e-01) | 0.95 [0.81 1.10] (4.68e-01) | 0.74 [0.50 1.09] (1.28e-01) | 1.09 [0.95 1.27] (2.23e-01) |
| **rs7912495** | 10:11676714:G:A | *ECHDC3* | 46.2% | 1.06 [1.05 1.08] (9.70e-19) | Bellenguez *et al.* (2022) | 46.1% | 1.17 [1.03 1.33] (1.47e-02) | 1.13 [1.01 1.25] (2.56e-02) | 1.24 [0.97 1.59] (8.33e-02) | 1.04 [0.94 1.15] (4.86e-01) |
| **rs10906923** | 10:15527599:C:A | *FAM171A1* | 30.6% | 0.93 [0.91 0.96] (1.35e-08) | Chang *et al.* (2017) | 32.4% | 1.03 [0.90 1.18] (6.87e-01) | 0.99 [0.89 1.11] (8.71e-01) | 0.83 [0.64 1.08] (1.68e-01) | 1.05 [0.94 1.17] (4.23e-01) |
| **rs7068231** | 10:60025170:T:G | *ANK3* | 40.3% | 0.95 [0.94 0.96] (3.30e-13) | Bellenguez *et al.* (2022) | 39.5% | 1.02 [0.89 1.17] (7.55e-01) | 1.01 [0.91 1.13] (8.30e-01) | 0.87 [0.68 1.12] (2.77e-01) | 0.96 [0.86 1.07] (4.59e-01) |
| **rs6586028** | 10:80494228:C:T | *TSPAN14* | 19.6% | 0.93 [0.91 0.94] (2.00e-19) | Bellenguez *et al.* (2022) | 19.4% | 0.90 [0.76 1.05] (1.85e-01) | 0.93 [0.81 1.06] (2.73e-01) | 1.08 [0.81 1.44] (6.02e-01) | 0.98 [0.86 1.12] (7.90e-01) |
| **rs6584063** | 10:96266650:G:A | *BLNK* | 4.3% | 0.89 [0.86 0.92] (6.70e-11) | Bellenguez *et al.* (2022) | 3.5% | 0.83 [0.60 1.15] (2.63e-01) | 0.80 [0.61 1.04] (9.85e-02) | 0.85 [0.46 1.58] (6.07e-01) | 1.07 [0.80 1.44] (6.51e-01) |
| **rs10748818** | 10:102255522:G:A | *GBF1* | 14.9% | 1.09 [1.05 1.11] (1.05e-09) | Nalls *et al.* (2019) | 14.5% | 0.90 [0.75 1.08] (2.70e-01) | 0.98 [0.85 1.13] (7.65e-01) | 0.94 [0.66 1.33] (7.12e-01) | 0.91 [0.79 1.06] (2.28e-01) |
| **rs117896735** | 10:119776815:A:G | *BAG3* | 1.2% | 1.65 [1.48 1.85] (2.23e-19) | Chang *et al.* (2017) | 1.7% | 1.62 [0.98 2.67] (5.93e-02) | 1.37 [0.89 2.12] (1.56e-01) | 1.03 [0.35 3.04] (9.56e-01) | 1.10 [0.75 1.60] (6.28e-01) |
| **rs7908662** | 10:122413396:G:A | *PLEKHA1* | 46.7% | 0.96 [0.95 0.97] (2.60e-09) | Bellenguez *et al.* (2022) | 47.4% | 1.00 [0.89 1.14] (9.41e-01) | 0.93 [0.83 1.03] (1.55e-01) | 0.95 [0.75 1.21] (6.73e-01) | 1.05 [0.95 1.17] (3.26e-01) |
| **rs7938782** | 11:10537230:G:A | *RNF141* | 12.2% | 0.92 [0.89 0.94] (2.12e-09) | Nalls *et al.* (2019) | 12.6% | 1.05 [0.87 1.27] (5.97e-01) | 0.98 [0.83 1.14] (7.55e-01) | 1.19 [0.85 1.68] (3.13e-01) | 1.09 [0.94 1.28] (2.54e-01) |
| **rs10437655** | 11:47370397:A:G | *CELF1/ SPI1* | 39.9% | 1.06 [1.04 1.07] (5.30e-14) | Bellenguez *et al.* (2022) | 39.8% | 1.12 [0.98 1.27] (8.43e-02) | 1.07 [0.96 1.18] (2.29e-01) | 1.01 [0.79 1.30] (9.17e-01) | 1.04 [0.93 1.15] (5.05e-01) |
| **rs1582763** | 11:60254475:A:G | *MS4A* | 37.1% | 0.91 [0.90 0.92] (3.70e-42) | Bellenguez *et al.* (2022) | 36.2% | 0.97 [0.86 1.11] (7.01e-01) | 0.97 [0.87 1.08] (5.32e-01) | 1.03 [0.80 1.31] (8.28e-01) | 1.05 [0.94 1.17] (3.64e-01) |
| **rs3793947** | 11:83833429:A:G | *DLG2* | 46.3% | 0.93 [0.91 0.95] (3.72e-09) | Chang *et al.* (2017) | 43.7% | 0.96 [0.84 1.09] (5.07e-01) | 1.01 [0.91 1.12] (9.24e-01) | 1.06 [0.83 1.34] (6.62e-01) | 0.92 [0.83 1.02] (1.33e-01) |
| **rs3851179** | 11:86157598:T:C | *PICALM* | 35.8% | 0.90 [0.89 0.92] (3.00e-48) | Bellenguez *et al.* (2022) | 35.2% | 0.91 [0.79 1.04] (1.51e-01) | 0.86 [0.77 0.96] (7.92e-03) | 1.05 [0.82 1.36] (6.99e-01) | 1.06 [0.95 1.18] (3.11e-01) |
| **rs74685827** | 11:121482368:G:T | *SORL1* | 1.9% | 1.19 [1.13 1.25] (2.80e-11) | Bellenguez *et al.* (2022) | 2.1% | 1.03 [0.65 1.64] (9.08e-01) | 1.14 [0.78 1.67] (5.08e-01) | 1.84 [0.88 3.86] (1.06e-01) | 0.90 [0.63 1.28] (5.45e-01) |
| **rs11218343** | 11:121564878:C:T | *SORL1* | 3.9% | 0.84 [0.81 0.87] (1.40e-21) | Bellenguez *et al.* (2022) | 3.6% | 1.02 [0.74 1.39] (9.15e-01) | 0.72 [0.55 0.95] (2.12e-02) | 1.72 [1.02 2.90] (4.36e-02) | 1.32 [1.00 1.73] (4.79e-02) |
| **rs329648** | 11:133895472:T:C | *MIR4697* | 32.7% | 1.09 [1.07 1.12] (1.11e-13) | Chang *et al.* (2017) | 35.0% | 0.96 [0.84 1.10] (5.66e-01) | 1.10 [0.98 1.23] (1.11e-01) | 1.03 [0.79 1.33] (8.33e-01) | 0.88 [0.79 0.99] (2.62e-02) |
| **rs76904798** | 12:40220632:T:C | *LRRK2* | 13.2% | 1.15 [1.12 1.19] (1.21e-19) | Chang *et al.* (2017) | 13.0% | 0.90 [0.75 1.08] (2.66e-01) | 0.89 [0.77 1.04] (1.38e-01) | 0.85 [0.59 1.22] (3.68e-01) | 0.98 [0.84 1.14] (8.10e-01) |
| **rs7134559** | 12:46025303:T:C | *SCAF11* | 40.4% | 0.95 [0.93 0.97] (3.96e-08) | Nalls *et al.* (2019) | 39.8% | 0.92 [0.81 1.05] (2.04e-01) | 0.93 [0.84 1.04] (2.05e-01) | 1.00 [0.79 1.27] (1.00e+00) | 0.98 [0.88 1.09] (7.35e-01) |
| **rs6489896** | 12:113281983:C:T | *TPCN1* | 7.6% | 1.08 [1.05 1.10] (1.80e-09) | Bellenguez *et al.* (2022) | 7.0% | 1.31 [1.01 1.69] (4.04e-02) | 1.22 [0.99 1.52] (6.46e-02) | 1.29 [0.80 2.10] (3.00e-01) | 1.00 [0.82 1.22] (9.80e-01) |
| **rs11060180** | 12:122819039:G:A | *OGFOD2* | 45.0% | 0.90 [0.88 0.92] (2.05e-20) | Chang *et al.* (2017) | 45.3% | 1.06 [0.94 1.20] (3.58e-01) | 1.06 [0.95 1.17] (2.83e-01) | 1.10 [0.87 1.40] (4.16e-01) | 0.97 [0.87 1.07] (5.40e-01) |

| | | | | | | | | | | |
|---|---|---|---|---|---|---|---|---|---|---|
| **rs11610045** | 12:132487182:A:G | *FBRSL1* | 49.0% | 1.06 [1.04 1.08] (1.77e-10) | Nalls *et al.* (2019) | 48.2% | 0.86 [0.76 0.98] (2.03e-02) | 0.91 [0.82 1.01] (7.42e-02) | 0.92 [0.71 1.18] (4.92e-01) | 0.97 [0.87 1.07] (5.27e-01) |
| **rs9568188** | 13:49353596:C:T | *CAB39L* | 26.0% | 0.94 [0.92 0.96] (1.15e-08) | Nalls *et al.* (2019) | 26.9% | 0.97 [0.84 1.12] (6.81e-01) | 1.01 [0.90 1.14] (8.43e-01) | 0.95 [0.73 1.25] (7.24e-01) | 0.94 [0.84 1.06] (3.26e-01) |
| **rs4771268** | 13:97212767:T:C | *MBNL2* | 23.0% | 1.07 [1.05 1.09] (1.45e-09) | Nalls *et al.* (2019) | 22.6% | 0.93 [0.79 1.08] (3.39e-01) | 1.02 [0.90 1.16] (7.04e-01) | 1.12 [0.84 1.48] (4.45e-01) | 0.91 [0.80 1.03] (1.36e-01) |
| **rs12147950** | 14:37520065:T:C | *MIPOL1* | 43.8% | 0.95 [0.93 0.97] (3.54e-08) | Nalls *et al.* (2019) | 43.2% | 1.07 [0.94 1.22] (3.22e-01) | 1.10 [0.99 1.22] (8.29e-02) | 1.29 [1.00 1.65] (4.59e-02) | 0.95 [0.86 1.06] (3.49e-01) |
| **rs17125924** | 14:52924962:G:A | *FERMT2* | 8.9% | 1.10 [1.07 1.12] (8.30e-16) | Bellenguez *et al.* (2022) | 9.0% | 1.27 [1.00 1.60] (4.95e-02) | 1.38 [1.14 1.67] (8.73e-04) | 1.17 [0.75 1.82] (4.79e-01) | 1.17 [0.75 1.82] (4.79e-01) |
| **rs11158026** | 14:54882151:T:C | *GCH1* | 30.7% | 0.91 [0.89 0.93] (4.30e-16) | Chang *et al.* (2017) | 33.2% | 0.95 [0.84 1.09] (4.89e-01) | 0.98 [0.88 1.09] (6.80e-01) | 0.94 [0.73 1.20] (6.02e-01) | 0.97 [0.87 1.08] (5.98e-01) |
| **rs1555399** | 14:67517653:A:T | *TMEM229B* | 45.6% | 0.92 [0.90 0.94] (9.61e-11) | Chang *et al.* (2017) | 48.5% | 1.11 [0.98 1.25] (1.17e-01) | 1.00 [0.91 1.11] (9.41e-01) | 0.91 [0.72 1.16] (4.37e-01) | 1.10 [1.00 1.22] (5.79e-02) |
| **rs3742785** | 14:74906331:C:A | *RPS6KL1* | 21.3% | 0.93 [0.91 0.95] (1.92e-09) | Nalls *et al.* (2019) | 21.6% | 1.12 [0.96 1.30] (1.55e-01) | 1.10 [0.97 1.24] (1.58e-01) | 1.11 [0.83 1.49] (4.67e-01) | 1.02 [0.90 1.15] (7.68e-01) |
| **rs8005172** | 14:88006268:T:C | *GALC* | 42.4% | 1.08 [1.05 1.10] (8.77e-11) | Chang *et al.* (2017) | 43.4% | 1.03 [0.91 1.17] (6.33e-01) | 0.96 [0.87 1.07] (4.70e-01) | 0.89 [0.70 1.14] (3.54e-01) | 1.06 [0.96 1.18] (2.42e-01) |
| **rs7401792** | 14:92464917:G:A | *SLC24A4/ RIN3* | 37.1% | 1.04 [1.02 1.05] (4.80e-08) | Bellenguez *et al.* (2022) | 36.8% | 1.03 [0.90 1.18] (6.87e-01) | 1.10 [0.99 1.23] (7.90e-02) | 0.92 [0.71 1.20] (5.41e-01) | 0.96 [0.86 1.07] (4.44e-01) |
| **rs12590654** | 14:92472511:A:G | *SLC24A4/ RIN3* | 32.8% | 0.93 [0.92 0.95] (4.20e-21) | Bellenguez *et al.* (2022) | 33.2% | 0.89 [0.78 1.02] (1.04e-01) | 1.00 [0.89 1.11] (9.49e-01) | 1.30 [1.00 1.67] (4.65e-02) | 0.94 [0.85 1.05] (2.94e-01) |
| **rs7157106** | 14:105761758:A:G | *IGH* | 36.0% | 1.05 [1.03 1.07] (2.00e-08) | Bellenguez *et al.* (2022) | 31.7% | 1.05 [0.92 1.21] (4.62e-01) | 1.10 [0.98 1.24] (9.96e-02) | 0.91 [0.69 1.20] (5.07e-01) | 0.97 [0.87 1.09] (6.05e-01) |
| **rs8025980** | 15:50701814:G:A | *SPPL2A* | 34.5% | 0.96 [0.94 0.97] (1.30e-08) | Bellenguez *et al.* (2022) | 35.1% | 0.99 [0.86 1.12] (8.27e-01) | 0.96 [0.86 1.07] (4.46e-01) | 0.96 [0.75 1.23] (7.56e-01) | 1.05 [0.95 1.17] (3.54e-01) |
| **rs602602** | 15:58764824:A:T | *ADAM10* | 28.0% | 0.94 [0.93 0.96] (2.10e-15) | Bellenguez *et al.* (2022) | 28.0% | 1.00 [0.87 1.16] (9.62e-01) | 1.00 [0.90 1.13] (9.40e-01) | 1.08 [0.82 1.43] (5.67e-01) | 0.97 [0.87 1.09] (6.14e-01) |
| **rs2414739** | 15:61701935:G:A | *VPS13C* | 29.2% | 0.91 [0.89 0.93] (3.94e-14) | Chang *et al.* (2017) | 26.5% | 0.98 [0.85 1.13] (7.48e-01) | 0.91 [0.81 1.03] (1.36e-01) | 0.87 [0.66 1.15] (3.43e-01) | 1.06 [0.94 1.19] (3.58e-01) |
| **rs117618017** | 15:63277703:T:C | *APH1B* | 14.4% | 1.11 [1.09 1.13] (2.20e-25) | Bellenguez *et al.* (2022) | 13.6% | 1.15 [0.95 1.39] (1.43e-01) | 1.20 [1.03 1.40] (2.00e-02) | 1.44 [1.02 2.02] (3.76e-02) | 0.96 [0.83 1.12] (6.04e-01) |
| **rs3848143** | 15:64131307:G:A | *SNX1* | 22.0% | 1.05 [1.04 1.07] (8.40e-11) | Bellenguez *et al.* (2022) | 22.1% | 1.04 [0.89 1.22] (5.97e-01) | 1.05 [0.92 1.19] (5.00e-01) | 0.94 [0.69 1.27] (6.68e-01) | 1.00 [0.88 1.13] (9.46e-01) |
| **rs12592898** | 15:78936857:A:G | *CTSH* | 13.3% | 0.94 [0.92 0.96] (4.20e-09) | Bellenguez *et al.* (2022) | 12.6% | 0.87 [0.72 1.06] (1.65e-01) | 0.90 [0.77 1.05] (1.74e-01) | 0.89 [0.63 1.28] (5.38e-01) | 0.94 [0.80 1.10] (4.36e-01) |
| **rs11343** | 16:19268142:T:G | *COQ7* | 45.4% | 1.07 [1.05 1.10] (9.13e-11) | Chang *et al.* (2017) | 44.4% | 1.06 [0.93 1.20] (3.98e-01) | 0.99 [0.89 1.10] (8.64e-01) | 0.94 [0.74 1.20] (6.37e-01) | 1.10 [0.99 1.22] (9.05e-02) |
| **rs2904880** | 16:28933075:C:G | *CD19* | 30.9% | 0.94 [0.92 0.96] (7.87e-10) | Nalls *et al.* (2019) | 31.6% | 1.00 [0.87 1.15] (9.70e-01) | 1.08 [0.97 1.21] (1.62e-01) | 1.02 [0.78 1.32] (8.95e-01) | 0.95 [0.85 1.06] (3.88e-01) |

| | | | | | | | | | | |
|---|---|---|---|---|---|---|---|---|---|---|
| **rs1140239** | 16:30010081:T:C | *DOC2A* | 37.9% | 0.94 [0.93 0.96] (2.60e-13) | Bellenguez *et al.* (2022) | 38.0% | 0.81 [0.71 0.93] (3.17e-03) | 0.90 [0.81 1.00] (5.45e-02) | 0.79 [0.61 1.02] (6.90e-02) | 0.96 [0.86 1.07] (4.73e-01) |
| **rs14235** | 16:31110472:A:G | *ZNF646/ KAT8* | 39.7% | 1.08 [1.06 1.10] (5.44e-12) | Chang *et al.* (2017) | 37.7% | 1.01 [0.88 1.16] (8.69e-01) | 1.05 [0.94 1.17] (3.90e-01) | 1.08 [0.84 1.39] (5.48e-01) | 0.98 [0.88 1.09] (6.70e-01) |
| **rs889555** | 16:31111250:T:C | *KAT8* | 28.1% | 0.95 [0.94 0.97] (2.00e-11) | Bellenguez *et al.* (2022) | 27.8% | 0.93 [0.81 1.08] (3.46e-01) | 0.91 [0.81 1.02] (1.12e-01) | 0.89 [0.67 1.17] (4.04e-01) | 1.00 [0.89 1.12] (9.71e-01) |
| **rs6500328** | 16:50702745:G:A | *NOD2* | 40.1% | 0.94 [0.93 0.96] (1.82e-09) | Nalls *et al.* (2019) | 41.3% | 1.13 [0.99 1.28] (6.83e-02) | 1.10 [0.99 1.23] (6.40e-02) | 0.89 [0.69 1.14] (3.61e-01) | 1.03 [0.93 1.14] (5.84e-01) |
| **rs4784227** | 16:52565276:T:C | *TOX3* | 26.5% | 1.09 [1.06 1.12] (9.75e-11) | Chang *et al.* (2017) | 24.7% | 0.96 [0.83 1.11] (5.91e-01) | 0.96 [0.85 1.08] (5.34e-01) | 0.73 [0.54 0.98] (3.72e-02) | 1.00 [0.89 1.13] (9.54e-01) |
| **rs4985556** | 16:70660097:A:C | *IL34* | 11.5% | 1.07 [1.05 1.09] (6.00e-10) | Bellenguez *et al.* (2022) | 11.5% | 1.21 [0.99 1.47] (6.04e-02) | 1.03 [0.87 1.22] (7.14e-01) | 0.92 [0.61 1.37] (6.74e-01) | 1.14 [0.97 1.33] (1.01e-01) |
| **rs450674** | 16:79574511:C:T | *MAF* | 37.3% | 0.96 [0.95 0.98] (3.20e-08) | Bellenguez *et al.* (2022) | 36.8% | 0.96 [0.85 1.10] (5.89e-01) | 0.87 [0.78 0.97] (1.01e-02) | 0.96 [0.75 1.24] (7.74e-01) | 1.09 [0.98 1.21] (1.29e-01) |
| **rs12446759** | 16:81739398:G:A | *PLCG2* | 40.3% | 0.95 [0.94 0.96] (1.20e-13) | Bellenguez *et al.* (2022) | 39.8% | 1.04 [0.91 1.19] (5.44e-01) | 0.96 [0.86 1.06] (4.08e-01) | 1.04 [0.81 1.33] (7.74e-01) | 1.07 [0.96 1.19] (2.14e-01) |
| **rs16941239** | 16:86420604:A:T | *FOXF1* | 2.9% | 1.13 [1.08 1.17] (1.30e-08) | Bellenguez *et al.* (2022) | 2.6% | 0.94 [0.63 1.40] (7.57e-01) | 0.91 [0.66 1.25] (5.63e-01) | 0.87 [0.40 1.92] (7.38e-01) | 1.07 [0.78 1.47] (6.92e-01) |
| **rs56407236** | 16:90103687:A:G | *PRDM7* | 6.9% | 1.11 [1.08 1.14] (6.50e-15) | Bellenguez *et al.* (2022) | 6.3% | 1.38 [1.06 1.79] (1.67e-02) | 1.06 [0.85 1.32] (6.08e-01) | 0.79 [0.46 1.35] (3.88e-01) | 1.22 [0.99 1.50] (5.80e-02) |
| **rs35048651** | 17:1728046:T:TGAG | *WDR81* | 21.4% | 1.06 [1.04 1.08] (7.70e-11) | Bellenguez *et al.* (2022) | 21.6% | 0.95 [0.81 1.10] (4.98e-01) | 0.97 [0.86 1.10] (6.29e-01) | 0.94 [0.70 1.26] (6.91e-01) | 0.97 [0.85 1.09] (5.83e-01) |
| **rs7225151** | 17:5233752:A:G | *SCIMP/ RABEP1* | 12.4% | 1.08 [1.05 1.10] (4.10e-13) | Bellenguez *et al.* (2022) | 12.1% | 1.00 [0.82 1.22] (9.97e-01) | 1.11 [0.95 1.31] (1.95e-01) | 1.19 [0.82 1.72] (3.68e-01) | 1.00 [0.86 1.17] (9.88e-01) |
| **rs12600861** | 17:7452302:C:A | *CHRNB1* | 35.2% | 1.05 [1.04 1.08] (1.01e-08) | Nalls *et al.* (2019) | 35.3% | 1.00 [0.87 1.14] (9.71e-01) | 0.97 [0.87 1.09] (6.32e-01) | 1.16 [0.90 1.48] (2.44e-01) | 1.01 [0.91 1.13] (8.46e-01) |
| **rs2242595** | 17:18156140:A:G | *MYO15A* | 11.2% | 0.94 [0.92 0.96] (1.10e-09) | Bellenguez *et al.* (2022) | 11.3% | 0.87 [0.71 1.06] (1.69e-01) | 0.93 [0.80 1.10] (4.01e-01) | 0.91 [0.61 1.34] (6.20e-01) | 0.91 [0.77 1.07] (2.71e-01) |
| **rs601999** | 17:42546140:T:C | *ATP6V0A1/ PSMC3IP/ TUBG2* | 30.1% | 1.08 (8.03e-09) | Chang *et al.* (2017) | 33.3% | 1.07 [0.93 1.23] (3.44e-01) | 1.05 [0.94 1.18] (3.76e-01) | 1.16 [0.90 1.51] (2.51e-01) | 1.01 [0.90 1.13] (9.22e-01) |
| **rs2269906** | 17:44216969:C:A | *UBTF* | 34.7% | 0.93 [0.92 0.96] (6.24e-10) | Nalls *et al.* (2019) | 35.9% | 0.97 [0.85 1.11] (6.71e-01) | 0.96 [0.87 1.07] (5.17e-01) | 0.89 [0.69 1.15] (3.68e-01) | 1.00 [0.90 1.11] (9.79e-01) |
| **rs5848** | 17:44352876:T:C | *GRN* | 28.9% | 1.07 [1.06 1.09] (2.40e-20) | Bellenguez *et al.* (2022) | 29.5% | 0.98 [0.86 1.13] (8.12e-01) | 0.95 [0.85 1.07] (4.00e-01) | 1.02 [0.79 1.32] (8.60e-01) | 1.01 [0.90 1.13] (8.45e-01) |
| **rs850738** | 17:44357262:G:A | *FAM171A2* | 39.4% | 1.08 [1.05 1.10] (1.29e-11) | Nalls *et al.* (2019) | 40.9% | 1.03 [0.91 1.17] (6.49e-01) | 0.96 [0.86 1.06] (3.91e-01) | 1.02 [0.81 1.30] (8.51e-01) | 1.06 [0.96 1.18] (2.57e-01) |
| **rs17649553** | 17:45917282:T:C | *ARHGAP27/ CRHR1/ SPPL2C/ MAPT/ STH/ KANSL1* | 23.2% | 0.78 [0.76 0.80] (1.26e-68) | Chang *et al.* (2017) | 21.2% | 0.89 [0.76 1.05] (1.62e-01) | 0.91 [0.80 1.03] (1.21e-01) | 0.82 [0.60 1.12] (2.06e-01) | 0.96 [0.84 1.08] (4.89e-01) |

| | | | | | | | | | | |
|---|---|---|---|---|---|---|---|---|---|---|
| **rs199515** | 17:46779275:G:C | *MAPT* | 21.9% | 0.94 [0.93 0.96] (9.30e-13) | Bellenguez *et al.* (2022) | 20.4% | 0.93 [0.79 1.09] (3.54e-01) | 0.92 [0.81 1.05] (2.00e-01) | 0.84 [0.61 1.16] (2.85e-01) | 0.96 [0.85 1.10] (5.74e-01) |
| **rs616338** | 17:49219935:T:C | *ABI3* | 1.2% | 1.32 [1.23 1.42] (2.80e-14) | Bellenguez *et al.* (2022) | 1.1% | 2.35 [1.02 5.41] (4.47e-02) | 1.86 [0.86 4.04] (1.16e-01) | 2.64 [0.53 13.00] (2.34e-01) | 1.29 [0.80 2.07] (2.89e-01) |
| **rs2526377** | 17:58332680:G:A | *TSPOAP1* | 44.5% | 0.95 [0.94 0.97] (1.60e-12) | Bellenguez *et al.* (2022) | 44.0% | 0.91 [0.81 1.04] (1.58e-01) | 0.94 [0.85 1.05] (2.62e-01) | 1.19 [0.94 1.51] (1.48e-01) | 0.95 [0.86 1.06] (3.77e-01) |
| **rs61169879** | 17:61840005:T:C | *BRIP1* | 16.4% | 1.09 [1.06 1.11] (9.28e-10) | Nalls *et al.* (2019) | 15.7% | 1.12 [0.94 1.33] (2.14e-01) | 1.13 [0.98 1.31] (9.02e-02) | 0.92 [0.65 1.31] (6.50e-01) | 0.95 [0.83 1.10] (5.02e-01) |
| **rs4277405** | 17:63471557:C:T | *ACE* | 38.4% | 0.94 [0.93 0.95] (8.80e-20) | Bellenguez *et al.* (2022) | 36.7% | 1.06 [0.93 1.21] (3.85e-01) | 1.06 [0.95 1.19] (2.75e-01) | 1.07 [0.84 1.38] (5.70e-01) | 0.98 [0.88 1.09] (7.13e-01) |
| **rs666463** | 17:78429399:T:A | *DNAH17* | 16.7% | 0.93 [0.90 0.95] (3.20e-09) | Nalls *et al.* (2019) | 16.6% | 1.00 [0.85 1.19] (9.75e-01) | 0.97 [0.85 1.12] (7.21e-01) | 1.01 [0.73 1.41] (9.46e-01) | 1.02 [0.89 1.16] (8.30e-01) |
| **rs8087969** | 18:51157219:G:T | *MEX3C* | 45.0% | 1.06 [1.04 1.08] (1.41e-08) | Nalls *et al.* (2019) | 44.8% | 1.03 [0.90 1.17] (6.90e-01) | 0.99 [0.89 1.10] (8.90e-01) | 0.91 [0.71 1.16] (4.42e-01) | 1.03 [0.93 1.14] (5.37e-01) |
| **rs12151021** | 19:1050875:A:G | *ABCA7* | 33.6% | 1.10 [1.09 1.12] (1.60e-37) | Bellenguez *et al.* (2022) | 33.3% | 1.23 [1.07 1.42] (3.69e-03) | 1.21 [1.08 1.35] (1.18e-03) | 1.01 [0.77 1.32] (9.35e-01) | 1.02 [0.92 1.14] (6.98e-01) |
| **rs149080927** | 19:1854254:G:GC | *KLF16* | 48.0% | 1.05 [1.04 1.07] (5.10e-10) | Bellenguez *et al.* (2022) | 47.2% | 1.03 [0.91 1.17] (5.97e-01) | 1.03 [0.93 1.14] (5.94e-01) | 1.15 [0.91 1.45] (2.33e-01) | 1.04 [0.93 1.15] (5.15e-01) |
| **rs62120679** | 19:2363321:T:C | *LSM7* | 32.4% | 1.08 [1.05 1.11] (6.64e-07) | Chang et *al.* (2017) | 30.2% | 0.92 [0.80 1.06] (2.48e-01) | 0.98 [0.88 1.10] (7.51e-01) | 1.25 [0.96 1.62] (9.53e-02) | 1.00 [0.89 1.12] (9.94e-01) |
| **rs9304690** | 19:49950060:T:C | *SIGLEC11* | 24.0% | 1.05 [1.03 1.07] (4.70e-09) | Bellenguez *et al.* (2022) | 25.0% | 1.19 [1.03 1.37] (1.87e-02) | 1.02 [0.90 1.15] (7.80e-01) | 1.05 [0.80 1.40] (7.12e-01) | 1.10 [0.98 1.23] (1.13e-01) |
| **rs587709** | 19:54267597:C:T | *LILRB2* | 32.5% | 1.05 [1.04 1.07] (3.60e-11) | Bellenguez *et al.* (2022) | 31.6% | 1.10 [0.96 1.26] (1.85e-01) | 1.05 [0.93 1.17] (4.43e-01) | 0.95 [0.73 1.24] (7.19e-01) | 1.05 [0.94 1.17] (4.34e-01) |
| **rs1358782** | 20:413334:A:G | *RBCK1* | 24.6% | 0.95 [0.94 0.97] (1.60e-08) | Bellenguez *et al.* (2022) | 23.7% | 1.01 [0.87 1.18] (8.87e-01) | 0.94 [0.83 1.07] (3.67e-01) | 0.93 [0.70 1.25] (6.47e-01) | 1.10 [0.97 1.24] (1.44e-01) |
| **rs8118008** | 20:3187520:G:A | *DDRGK1* | 40.4% | 1.07 [1.04 1.09] (1.99e-06) | Chang et *al.* (2017) | 39.3% | 1.05 [0.93 1.20] (4.22e-01) | 1.04 [0.93 1.15] (4.97e-01) | 0.92 [0.72 1.18] (4.96e-01) | 1.03 [0.93 1.14] (5.91e-01) |
| **rs77351827** | 20:6025395:T:C | *CRLS1* | 12.8% | 1.08 [1.05 1.11] (8.87e-09) | Nalls *et al.* (2019) | 11.9% | 0.89 [0.73 1.09] (2.51e-01) | 0.94 [0.80 1.10] (4.64e-01) | 0.94 [0.64 1.38] (7.55e-01) | 0.95 [0.81 1.12] (5.49e-01) |
| **rs6014724** | 20:56423488:G:A | *CASS4* | 9.0% | 0.89 [0.87 0.91] (4.10e-21) | Bellenguez *et al.* (2022) | 8.4% | 1.00 [0.80 1.24] (9.69e-01) | 0.90 [0.75 1.07] (2.30e-01) | 0.91 [0.58 1.40] (6.56e-01) | 1.05 [0.87 1.26] (6.26e-01) |
| **rs6742** | 20:63743088:T:C | *SLC2A4RG* | 22.1% | 0.95 [0.93 0.97] (2.60e-09) | Bellenguez *et al.* (2022) | 20.6% | 0.89 [0.76 1.04] (1.44e-01) | 0.93 [0.82 1.06] (2.87e-01) | 0.68 [0.48 0.94] (2.05e-02) | 0.95 [0.83 1.08] (4.26e-01) |
| **rs2154481** | 21:26101558:C:T | *APP* | 47.6% | 0.95 [0.94 0.97] (1.00e-12) | Bellenguez *et al.* (2022) | 46.9% | 1.03 [0.90 1.17] (6.85e-01) | 1.05 [0.94 1.17] (3.77e-01) | 1.22 [0.96 1.55] (1.11e-01) | 0.97 [0.88 1.08] (6.19e-01) |
| **rs2830489** | 21:26775872:T:C | *ADAMTS1* | 28.1% | 0.95 [0.94 0.97] (1.70e-10) | Bellenguez *et al.* (2022) | 28.0% | 0.95 [0.82 1.09] (4.30e-01) | 0.97 [0.87 1.09] (6.39e-01) | 1.08 [0.83 1.40] (5.77e-01) | 0.99 [0.89 1.11] (9.19e-01) |
| **rs2248244** | 21:37480059:A:G | *DYRK1A* | 28.3% | 1.07 [1.05 1.10] (2.74e-11) | Nalls *et al.* (2019) | 27.3% | 0.98 [0.85 1.14] (8.35e-01) | 0.99 [0.88 1.11] (8.33e-01) | 0.87 [0.66 1.16] (3.50e-01) | 0.98 [0.87 1.10] (7.21e-01) |
